# Does the use of prediction equations to correct self-reported height and weight improve obesity prevalance estimates? A pooled cross-sectional analysis of Health Survey for England data

**DOI:** 10.1101/2022.01.28.22270014

**Authors:** Shaun Scholes, Linda Ng Fat, Alison Moody, Jennifer S Mindell

## Abstract

**Objective:** Adults typically overestimate height and underestimate weight compared with directly measured values, and such misreporting varies by sociodemographic and health-related factors. Using self-reported and interviewer-measured height and weight, collected from the same participants, we aimed to develop a set of prediction equations to correct bias in self-reported height and weight, and assess whether this adjustment improved the accuracy of obesity prevalence estimates relative to those based only on self-report.

**Design:** Population-based cross-sectional study.

**Participants:** 38,942 participants aged 16+ (Health Survey for England 2011-16) with non-missing self-reported and interviewer-measured height and weight.

**Main outcome measures:** Comparisons between self-reported, interviewer-measured (gold standard) and corrected (based on prediction equations) body mass index (BMI: kg/m^2^) including (i) difference between means and obesity prevalence, and (ii) measures of agreement for BMI classification.

**Results:** On average, men overestimated height more than women (1.6 and 1.0cm, respectively; *p*<0.001), whilst women underestimated weight more than men (2.1 and 1.5kg, respectively; *p*<0.001). Underestimation of BMI was larger on average for women than for men (1.1 and 1.0kg/m^2^, respectively; *p*<0.001). Obesity prevalence based on self-reported BMI was 6.8 and 6.0 percentage points (pp) lower than that estimated using measured BMI for men and women, respectively. Corrected BMI (based on models containing all significant predictors of misreporting of height and weight) lowered underestimation of obesity to 0.8pp in both sexes and improved the sensitivity of being classified as obese over self-reported BMI by 15.0pp for men and 12.2pp for women. Results based on models using age alone as a predictor of misreporting were similar.

**Conclusions:** Compared with self-reported data, applying prediction equations improved the accuracy of obesity prevalence estimates and increased sensitivity of being classified as obese. Including additional sociodemographic variables does not add enough predictive power to justify the added complexity of including them in prediction equations.

**Strengths and limitations of the study:**

- The limitations of body mass index (BMI) calculated from self-reported values of height and weight are well known.
- Health examination surveys such as the Health Survey for England (HSE) enable study of reporting bias when they collect both self-report and directly measured data on height and weight from the same participants.
- This study used HSE 2011-16 data to derive a set of adjustments to self-reported height and weight based on linear regression models that estimated measured values of height and weight from self-reported values of height and weight, with additional corrections for sociodemographic and health-related factors predictive of misreporting.
- Corrected and measured BMI (the gold standard) were compared to quantify by how much obesity prevalence estimates were improved relative to those based on self-report data only.
- Prediction equations are specific to time, place, target population and methods of data collection. As such, these may not be applicable to surveys with more recent data, or different sociodemographic, health and self-reported anthropometric profiles.

## INTRODUCTION

A few large cross-sectional surveys in England and the United States (US) include only self-reports of height and weight, as direct measurement is not feasible due to factors such as cost. In lieu of direct measures, body mass index (BMI) derived from self-reported values of height and weight (hereafter, self-reported BMI) is sometimes used for research^1^ and regularly for estimating obesity prevalence at a sub-national level as part of monitoring efforts^2 3^. However, systematic literature reviews^4 5^ and epidemiologic studies^6-11^ have consistently shown that adults on average overestimate height and underestimate weight compared with measured values. Whilst such misreporting is typically moderate on average for continuous variables, self-reported BMI often results in a systematic (downward bias) misclassification of BMI categories. Misreporting of height and weight, plus the positive skewness of BMI distributions, result in significant underestimation of obesity prevalence^11^.

Misreporting of height and weight varies by sociodemographic factors, e.g. by sex^11^, age^12^, race/ethnicity^10^, and socioeconomic status^9^, and by health-related factors such as current smoking status^8^ and self-perceived health^8^. Younger women in particular underestimate weight (linked to social desirability bias)^13^, while older persons overestimate height (linked to reporting height measured earlier in life, prior to becoming shorter due to changes in bone and muscle)^6 12^. Misreporting of weight is greater in higher BMI categories^10 11 14-16^: the term “flat slope syndrome” in obesity epidemiology describes the systematic tendency for self-reported BMI (relative to measured BMI) to overestimate low values and underestimate high values^7 17 18^.

Health examination surveys (e.g. the Health Survey for England (HSE) and the US National Health and Nutrition Examination Survey (NHANES)) often collect both self-report and directly measured data on height and weight from the same participants, enabling study of self-reporting bias^10 11^. As such analyses have shown systematic patterns in misreporting, equations including variables predictive of misreporting have been developed for use with surveys collecting self-report but not measured height and weight^9 10 16^. In England, self-reported height and weight in the Active Lives Survey (ALS) datasets are adjusted by formulae based on HSE data to monitor excess weight (BMI ≥25kg/m^2^) at local authority level. For surveys with no direct measurements of height and weight, such adjustments are made in the expectation that corrected values of height and weight improve BMI classification sensitivities, and so can be used to estimate levels of excess weight and obesity more accurately, compared with reliance on self-report data alone^3 16^.

The HSE is the main data source for monitoring overweight and obesity in the general population in England. Annually since 1991, trained interviewers have measured participants ‘ height and weight. Self-reported height and weight were included in each survey year between 2011 and 2016. The present study aims to analyse HSE 2011-16 data to develop a set of equations to correct self-reported values of height and weight to more closely approximate measured values of height and weight. Should corrected BMI show an improvement over self-reported BMI, these equations could then be applied to (i) self-report data in HSE 2021 (where interviewer-measurement was not possible for a substantial portion of fieldwork due to Covid-19 pandemic precautions) and (ii) other interview-based surveys (e.g. ALS) to improve accuracy of obesity prevalence estimates. Our objectives were to (i) identify which variables are associated with misreporting (thereby meriting inclusion in prediction equations), and (ii) assess whether applying the chosen equations improved the classification of adults into BMI categories.

## METHODS

### Study design and participants

The present study used HSE data on adults (aged 16y+) from all survey years between 2011 and 2016, when both self-reported and measured height and weight were collected. The HSE is a cross-sectional, general population survey of individuals living in private households, with a new sample each year randomly selected by address^19^. Data collection occurs throughout the year. The first stage is a health interview, including questions about sociodemographic factors, diagnosed health conditions, self-rated health, health-related lifestyle behaviours, and direct measurements of – and in 2011-2016, self-reported - height and weight. The second stage is a nurse-visit, including biophysical measurements. Interviews and nurse visits take place in the participants ‘ own homes. All adults in selected households were eligible (maximum 10); the percentage of eligible households participating ranged from 66% in 2011 to 59% in 2016. Participants gave verbal consent to be interviewed, visited by a nurse, and to have anthropometric measurements taken. Research ethics approval was obtained from relevant committees.

Self-reported height and weight were collected with the questions: “*How tall are you without shoes?*” and “*How much do you weigh without clothes and shoes*?” Height was reported in either metres or feet and inches; weight was reported in kilogrammes (kg) or stones and pounds. Height was measured using a portable stadiometer with a sliding head plate, a base plate and connecting rods marked with a measuring scale. Participants were asked to remove their shoes. One measurement (to the nearest even millimetre) was taken, with participants ‘ stretching to the maximum height and the head positioned in the Frankfort plane. For those not pregnant, a single weight measurement (to the nearest 100g; maximum 200kg) was recorded using Class III Seca scales; participants were asked to remove their shoes and any bulky clothing or heavy items in pockets, etc. No adjustment was made for the weight of clothing. Participants unable to stand or unsteady on their feet were not measured. Those who weighed 200+kg were asked for their estimated weight because the scales are inaccurate above this level: these two cases were included herein (measured weight set equal to reported weight) to avoid underestimating weight in the upper-tail. Participants were assigned missing values if they were considered by the interviewer to have unreliable measurements, e.g. those who were too stooped or wore excessive clothing. Participants were not told at the time of interview that their height and weight would be measured; however, given their informed consent, it is likely that they might have anticipated being measured subsequent to their report.

### Analytical sample

All participants (n=49,817) were asked their height and weight soon after starting the interview, and the measurements took place near the end. As our aim was to study self-reporting bias, the analytical sample was limited to n=38,942 participants with non-missing self-reported and measured height and weight. The participants excluded was as follows: pregnant (n=471), missing self-report and measured data (n=1001), missing self-report but not measured data (n=2550), and missing measured but not self-report data (n=6853). Missing self-report and measured data on height and weight were primarily due to “don ‘t knows” and refusals, respectively.

### Statistical analyses

#### 1. Descriptive analysis: comparing self-reported and measured data

We decided *a priori* to conduct sex-specific analyses due to documented differences in reporting^6 8-10 14 20^ and the sexual dimorphisms in height, weight and adiposity^21^. Initial analyses showed no linear trend in misreporting over the six-year period in either sex (Supplementary Data Table S1): pooled data was therefore used for subsequent analysis.

Among complete cases (n=38,942), self-reported and measured mean height, weight and BMI were calculated with 95% confidence intervals (CIs). To compare across distributions^22^, we computed values at the 5^th^, 10^th^, 25^th^, 50^th^, 75^th^, 90^th^, and 95^th^ percentiles^11^. BMI was calculated as weight in kg divided by height in metres squared, and the World Health Organization classification was used to classify participants into five categories: underweight (BMI:<18.5kg/m^2^); normal weight (18.5-24.9kg/m^2^); overweight (25.0-29.9kg/m^2^); obesity grades I and II (30.0-39.9kg/m^2^) and obesity grade III (≥40kg/m^2^)^23^. Participants were also classified according to the binary categories of (i) overweight (including obesity), also described herein as excess weight (≥25kg/m^2^), and (ii) obesity (≥30kg/m^2^). These definitions were applied to all participants as adults are defined in the HSE series as aged 16y+.

To compare self-report and measured data, we first calculated the difference (self-reported minus measured) between means (height, weight and BMI) and between prevalence (overweight (including obesity), and obesity). The degree of individual variability in the difference was summarised by the standard deviation (SD)^4 6^. Bland-Altman limits of agreement (LOA) were calculated for BMI. Secondly, we cross-tabulated self-reported and measured BMI categories. Using measured BMI as gold standard, we calculated estimates of sensitivity (the percent of true positives) and specificity (the percent of true negatives) to quantify the classification accuracy of self-reported BMI. Cohen ‘s Kappa (κ) statistic was also used to assess the degree of agreement after accounting for agreement at random^13^.

#### 2. Developing prediction equations in our main analysis

Linear regression modelling was used to develop prediction equations that estimated measured values of height and weight from self-reported values of height and weight, with appropriate adjustments for any variables independently associated with misreporting^8-10 16 24^. This involved three main steps.

#### Step 1: Predictors of misreporting

First, as in other studies^6 9 20 25^, participants with an absolute difference (self-reported minus measured) ≥4SD from the mean were considered outliers (with possible unrealistic reported values): these were excluded (height: n=189; weight: n=276) to avoid potentially undue influence on the equations. For those remaining (n=38,477), separately for height and weight, linear regression modelling was used to identify which variables were independently predictive of the difference between self-reported and measured values. Continuous variables for self-reported height and weight were entered as linear, quadratic, and cubic terms to allow for possible non-linearity^10^; higher power terms were excluded if the slope was not statistically different from zero (*p*>0.05). Independent variables included age group (16-17y, 18-19y, and in 5-yr intervals up to 85y+); ethnic group; Government Office Region; cigarette smoking status (current, ex-, never-smoker); general health (very good/good, fair, bad/very bad); presence of a limiting longstanding illness; and two indicators of socioeconomic status: (i) highest educational qualification (university degree or equivalent, A level/diploma, O level/GCSE/vocational equivalent, or none) and (ii) Index of Multiple Deprivation (IMD) quintile, a small-area based measure of deprivation (least deprived to most deprived). To maximise sample sizes, missing values on independent variables were assigned to a separate category (n≥30) or the modal category (n<30). These variables were chosen based on a review of the literature and data availability (collected in HSE 2011-16 main interview). Based on joint Wald tests, variables statistically significant as a whole (*p*<0.05) were candidates for inclusion in prediction equations.

#### Step 2: Deriving the prediction equations

Secondly, as in other studies^9 20^, the sample was randomly split into a training (hereafter, split-sample A) and testing (split-sample B) dataset using a 70:30 ratio. Split-sample A (n=27,035) was used for model-fitting and refinement, and parameter estimation (prediction equations). Split-sample B (n=11,442) provided an independent assessment of predictive accuracy of the equations.

To develop the prediction equations via linear regression modelling, measured values of height and weight were the dependent variables^8^, and self-reported values of height and weight, age group, and any other variables significantly associated with misreporting (from Step 1) were the independent variables. In a final refinement step, only significant variables (*p*<0.05) were retained (hereafter, full models) for reasons of parsimony (i.e. achieved similar goodness-of-fit with as few predictors as possible). We also fitted models containing age group only as a predictor of misreporting (hereafter, reduced models): such equations may be particularly useful for researchers using surveys that do not contain all the independent variables retained in the full models.

#### Step 3: Assessing the predictive accuracy of the equations

Thirdly, the prediction equations generated from split-sample A were applied to split-sample B. Corrected BMI was derived using the predicted values of height and weight. Descriptive statistics (means for continuous variables and percentages for BMI categories) were used to compare self-reported, measured, and corrected values. Using measured BMI as gold standard, estimates of sensitivity and specificity were calculated to quantify by how much corrected BMI improved BMI classification. Finally, to investigate the presence of any systematic error in *self-reported* BMI (i.e. the “flat slope syndrome” mentioned earlier), we fitted a linear regression model in which the difference between self-reported and measured BMI was the dependent variable and measured BMI was the independent variable. To investigate any such error in *corrected* BMI, we fitted a linear regression model in which the difference between corrected and measured BMI was the dependent variable and measured BMI was the independent variable^16^. For each case, a significantly negative slope for BMI would indicate the tendency for BMI underestimation to increase as measured BMI increases.

### Sensitivity analysis

We also examined predictive accuracy of equations derived using a simpler approach, based on linear regression models with the measured values of height and weight as the dependent variable and the self-reported values of height and weight plus age (in single-year bands but trimmed at 90y) as independent variables^10 26^. Linear, quadratic and cubic terms were entered for self-reported height and weight, and linear and quadratic terms were entered for age. Each term was retained in the model (thereby included in the equation) irrespective of statistical significance to allow for possible associations in future datasets^10^. This approach (implemented using formulae based on HSE 2012-14 data) is currently used to correct self-reported values of height and weight in the ALS for monitoring levels of excess weight across English local authorities.

All analyses accounted for the complex survey design, incorporating survey non-response weights and geographical clustering. Statistical significance was set at *p*<0.05 for two-tailed tests, with no adjustment for multiple comparisons. HSE datasets are available via the UK Data Service (**www.ukdataservice.ac.uk**)^27-32^ and are subject to an end user license agreement. Dataset preparation was performed in SPSS v24.0 (IBM Corp., Armonk, New York); analysis was performed in Stata v17.1 (StataCorp LLC, College Station, Texas).

### Patient and public involvement

Patients or the public were not involved in the design, or conduct, or reporting, or dissemination plans of our research (which involves secondary analysis of existing data). The project was shaped by discussions with the HSE Steering Group, including representatives from various national government agencies and local authorities.

## RESULTS

### Comparing self-reported and measured height, weight and BMI

To compare self-reported and measured data, Table 1 presents the difference between means (height, weight, BMI) and prevalence (BMI categories). On average, men overestimated height more than women (difference: 1.6 and 1.0cm, respectively; *p*<0.001 for sex difference), whilst women underestimated weight more than men (difference: 2.1 and 1.5kg; *p*<0.001). Underestimation of BMI was slightly larger on average for women than for men (1.1 and 1.0kg/m^2^; *p*<0.001). About three-quarters of adults had self-reported BMI values within 2 units of measured BMI (data not shown); however, underestimation of BMI was greater in higher BMI categories (Supplementary Data Table S2). The Bland-Altman LOA show the range within which approximately 95% of the differences between self-reported and measured BMI would be expected to fall^33^. The LOA for self-reported BMI was in the range of -4.6 to 2.5 BMI units for men and from -5.0 to 2.7 units for women (unweighted data; Supplementary Data Figure S1). For both sexes, the highest percentiles of BMI were lower for self-reported than for measured data, indicating more compressed distributions (Supplementary Data Table S3; Figure S2). The prevalence of overweight (including obesity) was 8.0 and 9.0 percentage points (pp) lower for self-reported than measured data for men and women, respectively. The equivalent figures for obesity were 6.8 and 6.0pp.

**Table 1.**
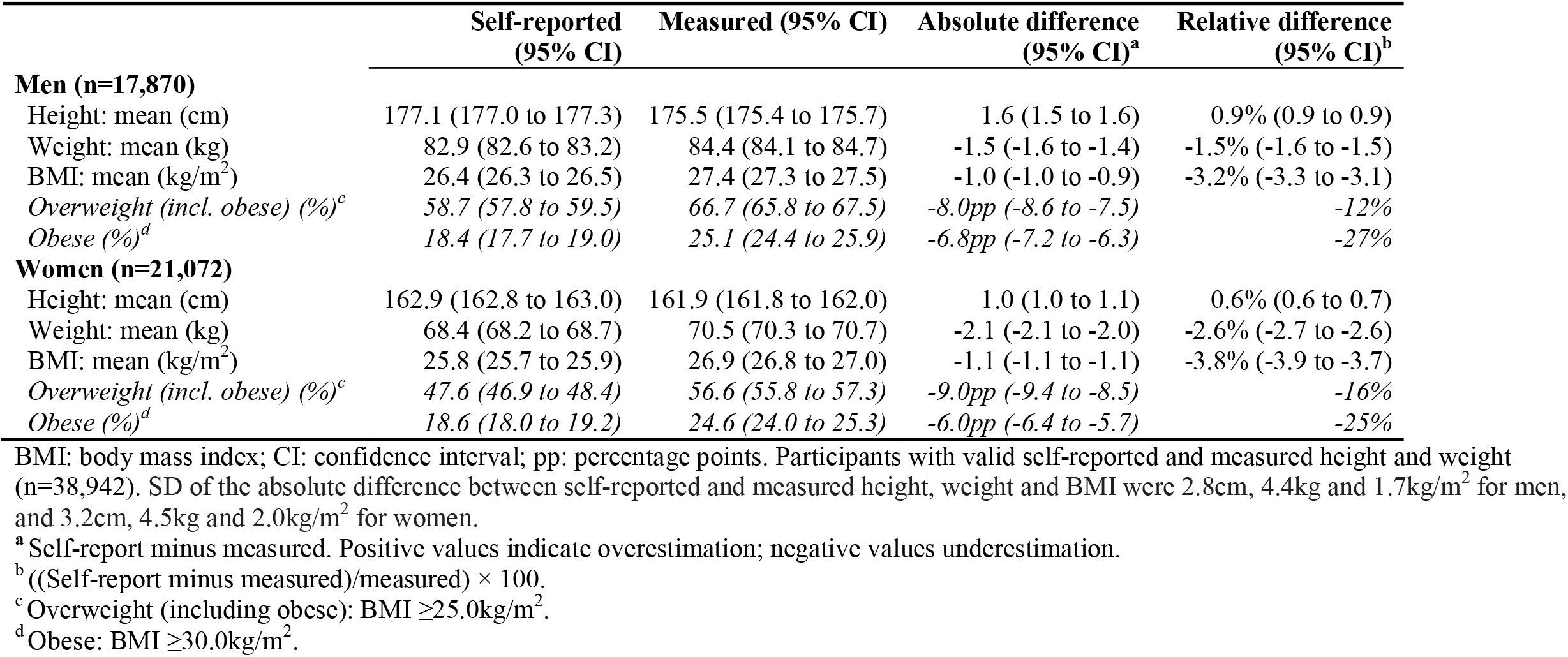
Means and differences in means for self-reported and measured height, weight and BMI by sex.

Using measured BMI as gold standard, 77% of men and 78% of women were correctly classified using self-reported BMI; the sensitivity of the self-reported obese category was 69% for men and 72% for women (Table 2). Of those classified as overweight (25.0-29.9kg/m^2^) based on self-reported BMI, 19% of men and 22% of women were classified as obese based on measured BMI.

**Table 2.** Cross-tabulation of measured and self-reported BMI categories by sex.

| Self-reported BMI categories | Measured BMI categories |  |  |  |  |
| --- | --- | --- | --- | --- | --- |
|  | Under-weight | Normal | Overweight | Obese I & II | Obese III |
|  | N (%) | N (%) | N (%) | N (%) | N (%) |
| **Men (n=17,870)** |  |  |  |  |  |
| Underweight | 209 (69.2) | 167 (2.7) | 4 (0.0) | 2 (0.0) | 0 (0.0) |
| Normal | 91 (30.1) | 5704 (91.0) | 1927 (23.5) | 49 (1.1) | 0 (0.0) |
| Overweight | 2 (0.7) | 390 (6.2) | 6081 (74.2) | 1475 (32.0) | 5 (1.4) |
| Obese I & II | 0 (0.0) | 7 (0.1) | 184 (2.2) | 3051 (66.2) | 178 (50.6) |
| Obese III | 0 (0.0) | 1 (0.0) | 1 (0.0) | 32 (0.7) | 168 (48.0) |
| Sensitivity | 69% | 91% | 74% | 66% | 48% |
| Specificity | 99% | 85% | 84% | 98% | 100% |
| κ | 0.66 |  |  |  |  |
| **Women (n=21,072)** |  |  |  |  |  |
| Underweight | 303 (73.7) | 292 (3.6) | 8 (0.1) | 0 (0.0) | 0 (0.0) |
| Normal | 106 (25.8) | 7524 (93.5) | 1886 (30.3) | 79 (1.9) | 5 (0.7) |
| Overweight | 2 (0.5) | 223 (2.8) | 4176 (67.1) | 1244 (29.9) | 10 (1.5) |
| Obese I & II | 0 (0.0) | 7 (0.1) | 152 (2.4) | 2796 (67.1) | 272 (43.3) |
| Obese III | 0 (0.0) | 1 (0.0) | 2 (0.0) | 47 (1.1) | 342 (54.5) |
| Sensitivity | 74% | 93% | 67% | 67% | 54% |
| Specificity | 98% | 82% | 89% | 97% | 100% |
| κ | 0.67 |  |  |  |  |

BMI: body mass index. Cell counts are weighted (rounded); estimates are column percentages. Participants with valid self-reported and measured height and weight (n=38,942). Shaded cells indicate those who were classified in the same category of BMI based on self-reported and measured height and weight. Underweight: BMI <18.5kg/m<sup>2</sup>; Normal: ≥18.5 – 24.9kg/m<sup>2</sup>; Overweight: ≥25.0 – 29.9kg/m<sup>2</sup>; Obese I & II: ≥30.0 – 39.9kg/m<sup>2</sup>; Obese III: ≥40.0kg/m<sup>2</sup>. κ (Cohen's kappa statistic)

### Predicting measured height from self-reported height and other variables (split-sample A)

Among men, based on multivariable linear regression analysis (dependent variable: self-reported minus measured height), older age, lower educational status (O level or no qualifications vs. having a degree), being Asian (vs. White), living in the North East, the North West and the West Midlands (vs. the South East), and reporting bad/very bad general health (vs. good/very good) were associated with greater overestimation of height. For women, older age, lower educational status (O level or no qualifications vs. having a degree), living in the North West (vs. the South East), living in the most (vs. least) deprived areas, and being in the Black, Asian or mixed ethnic groups (vs. White) were associated with greater overestimation of height (Supplementary Data Table S4).

The regression coefficients (prediction equations) for the aforementioned variables based on the models with measured height as the dependent variable correct self-reported height upwards (positive signs) or downwards (negative signs) as appropriate (Table S4): e.g., compared with those in the White group, the predicted measured height from the self-reported height of participants in the Asian group is corrected downwards by 0.61cm (men) and 1.78cm (women). The reduced models contained age group only as a predictor of misreporting. The R-squared (proportion of variance in measured height explained by the independent variables) for both the full and reduced models were similar for men (88.6% and 88.5%, respectively) and for women (87.7% and 87.1%).

### Predicting measured weight from self-reported weight and other variables (split-sample A)

For men, being an ex-regular or never (vs. current) smoker were associated with greater underestimation of weight; lower educational status (O level or no qualifications vs. having a degree) was associated with lower underestimation. For women, being an ex-regular or never (vs. current) smoker and being in the Black (vs. White) ethnic group were associated with greater underestimation of weight (Supplementary Data Table S5). Table S5 shows the prediction equations for deriving corrected values of weight: e.g. compared with current smokers, the predicted measured weight from the self-reported weight of never smokers was adjusted upwards by 0.65kg (men) and 0.18kg (women). The R-squared for both the full and reduced models were similar for men (94.3% for both models) and for women (95.1% for both models).

### Assessing the predictive accuracy of the equations (split-sample B)

The prediction equations were applied to split-sample B to generate corrected values. Table 3 shows the means (height, weight, BMI) and percentages (BMI categories) for the self-reported, measured, and corrected values. In each case, corrected estimates were closer than self-reported estimates to measured estimates, and the difference in means between corrected and measured values were not significantly different from zero, with the exception of height for women (e.g. measured height was underestimated on average by 0.1cm in the full model (*P*=0.003; data not shown)). The LOA for corrected BMI was in the range of -2.9 to 2.9 BMI units for men and from -3.0 to 3.0 units for women (unweighted data; Supplementary Data Figure S3). For each set of data, Figure 1 plots mean height and weight by sex and age group.

**Table 3.**
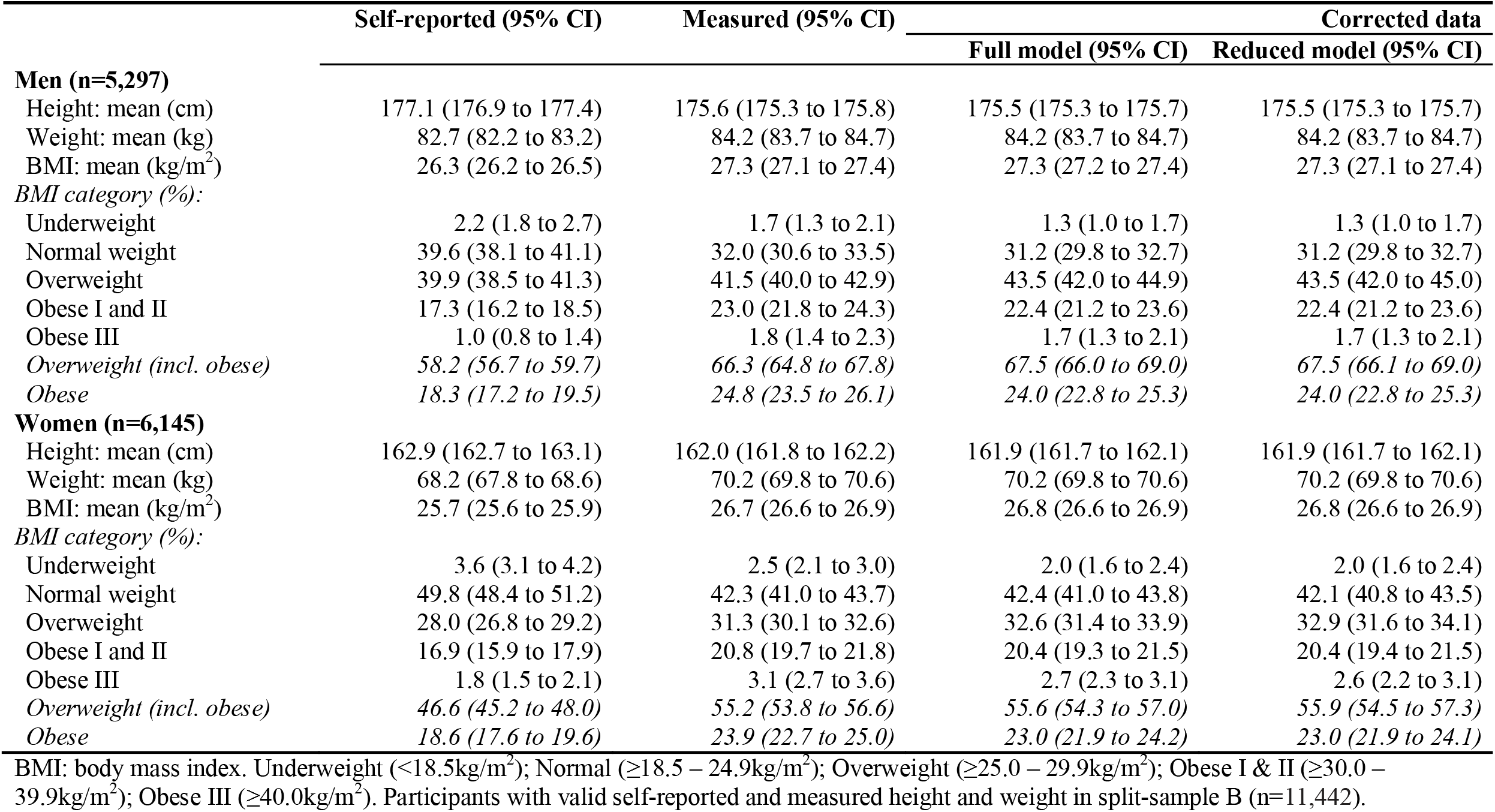
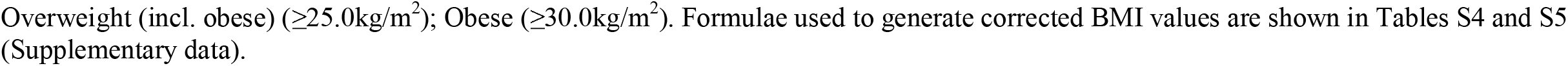
Mean height, weight and BMI for self-reported, measured and corrected data by sex (split-sample B)

|  | Self-reported (95% CI) | Measured (95% CI) | Corrected data |  |
| --- | --- | --- | --- | --- |
|  |  |  | Full model (95% CI) | Reduced model (95% CI) |
| **Men (n=5,297)** |  |  |  |  |
| Height: mean (cm) | 177.1 (176.9 to 177.4) | 175.6 (175.3 to 175.8) | 175.5 (175.3 to 175.7) | 175.5 (175.3 to 175.7) |
| Weight: mean (kg) | 82.7 (82.2 to 83.2) | 84.2 (83.7 to 84.7) | 84.2 (83.7 to 84.7) | 84.2 (83.7 to 84.7) |
| BMI: mean (kg/m<sup>2</sup>) | 26.3 (26.2 to 26.5) | 27.3 (27.1 to 27.4) | 27.3 (27.2 to 27.4) | 27.3 (27.1 to 27.4) |
| *BMI category (%):* |  |  |  |  |
| Underweight | 2.2 (1.8 to 2.7) | 1.7 (1.3 to 2.1) | 1.3 (1.0 to 1.7) | 1.3 (1.0 to 1.7) |
| Normal weight | 39.6 (38.1 to 41.1) | 32.0 (30.6 to 33.5) | 31.2 (29.8 to 32.7) | 31.2 (29.8 to 32.7) |
| Overweight | 39.9 (38.5 to 41.3) | 41.5 (40.0 to 42.9) | 43.5 (42.0 to 44.9) | 43.5 (42.0 to 45.0) |
| Obese I and II | 17.3 (16.2 to 18.5) | 23.0 (21.8 to 24.3) | 22.4 (21.2 to 23.6) | 22.4 (21.2 to 23.6) |
| Obese III | 1.0 (0.8 to 1.4) | 1.8 (1.4 to 2.3) | 1.7 (1.3 to 2.1) | 1.7 (1.3 to 2.1) |
| *Overweight (incl. obese)* | *58.2 (56.7 to 59.7)* | *66.3 (64.8 to 67.8)* | *67.5 (66.0 to 69.0)* | *67.5 (66.1 to 69.0)* |
| *Obese* | *18.3 (17.2 to 19.5)* | *24.8 (23.5 to 26.1)* | *24.0 (22.8 to 25.3)* | *24.0 (22.8 to 25.3)* |
| **Women (n=6,145)** |  |  |  |  |
| Height: mean (cm) | 162.9 (162.7 to 163.1) | 162.0 (161.8 to 162.2) | 161.9 (161.7 to 162.1) | 161.9 (161.7 to 162.1) |
| Weight: mean (kg) | 68.2 (67.8 to 68.6) | 70.2 (69.8 to 70.6) | 70.2 (69.8 to 70.6) | 70.2 (69.8 to 70.6) |
| BMI: mean (kg/m<sup>2</sup>) | 25.7 (25.6 to 25.9) | 26.7 (26.6 to 26.9) | 26.8 (26.6 to 26.9) | 26.8 (26.6 to 26.9) |
| *BMI category (%):* |  |  |  |  |
| Underweight | 3.6 (3.1 to 4.2) | 2.5 (2.1 to 3.0) | 2.0 (1.6 to 2.4) | 2.0 (1.6 to 2.4) |
| Normal weight | 49.8 (48.4 to 51.2) | 42.3 (41.0 to 43.7) | 42.4 (41.0 to 43.8) | 42.1 (40.8 to 43.5) |
| Overweight | 28.0 (26.8 to 29.2) | 31.3 (30.1 to 32.6) | 32.6 (31.4 to 33.9) | 32.9 (31.6 to 34.1) |
| Obese I and II | 16.9 (15.9 to 17.9) | 20.8 (19.7 to 21.8) | 20.4 (19.3 to 21.5) | 20.4 (19.4 to 21.5) |
| Obese III | 1.8 (1.5 to 2.1) | 3.1 (2.7 to 3.6) | 2.7 (2.3 to 3.1) | 2.6 (2.2 to 3.1) |
| *Overweight (incl. obese)* | *46.6 (45.2 to 48.0)* | *55.2 (53.8 to 56.6)* | *55.6 (54.3 to 57.0)* | *55.9 (54.5 to 57.3)* |
| *Obese* | *18.6 (17.6 to 19.6)* | *23.9 (22.7 to 25.0)* | *23.0 (21.9 to 24.2)* | *23.0 (21.9 to 24.1)* |

BMI: body mass index. Underweight (<18.5kg/m<sup>2</sup>); Normal (≥18.5 – 24.9kg/m<sup>2</sup>); Overweight (≥25.0 – 29.9kg/m<sup>2</sup>); Obese I & II (≥30.0 – 39.9kg/m<sup>2</sup>); Obese III (≥40.0kg/m<sup>2</sup>). Participants with valid self-reported and measured height and weight in split-sample B (n=11,442).
Overweight (incl. obese) ( $\geq 25.0 \text{ kg/m}^2$ ); Obese ( $\geq 30.0 \text{ kg/m}^2$ ). Formulae used to generate corrected BMI values are shown in Tables S4 and S5 (Supplementary data).

**Figure 1.** Self-reported, measured and corrected height and weight by age-group and sex (men: left-panel; women : right-panel).

Compared with measured BMI, obesity prevalence based on self-reported BMI was 6.5 and 5.2pp lower for men and women, respectively. The corresponding value for corrected BMI (full models) was 0.8pp for both sexes (Table 3; Figure 2). The prevalence of overweight (including obesity) was slightly overestimated (1.2% men; 0.4% women).

**Figure 2.** Difference in obesity prevalence across population subgroups by sex (men: left-panel; women : right-panel). Negative values indicate underestimation; positive values overestimation.

Table 4 shows the cross-tabulation of corrected (full models) and measured BMI categories. 83% of men and 84% of women were correctly classified. Sensitivity values of the obesity category based on self-reported BMI were 71% and 75% for men and women, respectively (data not shown). The sensitivity of obesity based on corrected BMI increased to 86% and 87% for men and women, respectively, an absolute improvement over self-reported data by 15.0 and 12.2pp. The corresponding sensitivities based on reduced models were similar (86% both sexes: data not shown). In contrast, sensitivity values in the normal weight category (18.5-24.9kg/m^2^) were higher for self-reported (91% men; 94% women) than corrected BMI (83% men; 89% women).

**Table 4.** Cross-tabulation of measured and corrected BMI categories by sex (split-sample B)

| Corrected BMI categories (full model) | Measured BMI categories |  |  |  |  |
| --- | --- | --- | --- | --- | --- |
|  | Under-weight | Normal | Overweight | Obese I & II | Obese III |
|  | N (%) | N (%) | N (%) | N (%) | N (%) |
| **Men (n=5,297):** |  |  |  |  |  |
| Underweight | 55 (56.0) | 23 (1.2) | 0 (0.0) | 0 (0.0) | 0 (0.0) |
| Normal | 43 (44.1) | 1558 (82.8) | 232 (9.5) | 1 (0.1) | 0 (0.0) |
| Overweight | 0 (0.7) | 302 (16.0) | 2042 (83.8) | 209 (15.5) | 0 (0.0) |
| Obese I & II | 0 (0.0) | 0 (0.1) | 163 (6.7) | 1120 (82.8) | 31 (29.0) |
| Obese III | 0 (0.0) | 0 (0.0) | 0 (0.0) | 23 (1.7) | 75 (71.0) |
| Sensitivity | 56% | 83% | 84% | 83% | 71% |
| Specificity | 100% | 93% | 85% | 96% | 100% |
| $\kappa$ | 0.74 | | | | |
| **Women (n=6,145):** |  |  |  |  |  |
| Underweight | 84 (60.4) | 27 (1.1) | 0 (0.0) | 0 (0.0) | 0 (0.0) |
| Normal | 55 (39.6) | 2129 (88.7) | 216 (12.1) | 3 (0.0) | 0 (0.0) |
| Overweight | 0 (0.0) | 241 (10.1) | 1432 (80.6) | 175 (14.9) | 0 (0.0) |
| Obese I & II | 0 (0.0) | 2 (0.0) | 129 (7.3) | 975 (82.8) | 49 (27.7) |
| Obese III | 0 (0.0) | 0 (0.0) | 0 (0.0) | 24 (2.1) | 127 (72.3) |
| Sensitivity | 60% | 89% | 81% | 83% | 72% |
| Specificity | 100% | 92% | 89% | 96% | 100% |
| $\kappa$ | 0.76 | | | | |

Cell counts are weighted (rounded); estimates are column percentages. BMI: body mass index. Underweight: BMI <18.5kg/m<sup>2</sup>; Normal: ≥18.5 – 24.9kg/m<sup>2</sup>; Overweight: ≥25.0 – 29.9kg/m<sup>2</sup>; Obese I & II: ≥30.0 – 39.9kg/m<sup>2</sup>; Obese III: ≥40.0kg/m<sup>2</sup>. Participants with valid self-reported and measured height and weight in split-sample B (n=11,442). $\kappa$ (Cohen's Kappa). Formulae used to generate corrected BMI values are shown in Tables S4 and S5 (Supplementary data).

The results of linear regression analyses in which measured BMI was the predictor of error are shown in supplementary data (Table S6). The significantly negative slope for BMI (p<0.001 for both sexes) indicates that the differences between corrected and measured BMI showed a systematic, though smaller, bias in relation to measured BMI.

### Sensitivity analyses

The fitted regression equations describing the relationship between self-reported and measured height and weight with age as the single continuous predictor of misreporting are shown as supplementary data (Tables S7-S8; Figure S4 plots mean height and weight by sex and age group). The difference in means between the corrected and measured values were not significantly different from zero with the exception of height for women; the sensitivity estimates for this simpler method yielded similar values to the equations described above (overweight (including obesity): 94% men and 93% women; obesity: 85% men and 86% women; Table S9).

## DISCUSSION

Using pooled HSE 2011-16 data containing self-reported and measured height and weight, we developed two sets of prediction equations that can be easily used to correct for biases in self-reported BMI. Although not perfectly predictive of measured BMI, corrected BMI performed better than self-reported BMI in more closely approximating obesity prevalence based on measured BMI. Applying corrected values also increased sensitivity of the obesity category. Using measured BMI as gold standard, the sensitivity of obesity for the full model-corrected BMI was estimated as 86% and 87% for men and women, respectively, an improvement in absolute terms over self-reported BMI by 15.0 and 12.2pp.

### Misreporting

In agreement with other studies^11^, the present study showed that mean height was overestimated by self-report relative to measured height, and that mean weight was underestimated, resulting in a net underestimation of mean BMI. Our estimates for the difference between means in height, weight and BMI were within the range shown by systematic literature reviews^4 5^. In agreement with other studies^9 11^, we found that women underestimated weight more than men but that men overestimated height more than women. As reported elsewhere^34^, we do not know whether the differences between self-reported and measured data arise due to participants ‘ lack of knowledge about their current height and weight or whether it is due to misreporting of information that is accurately known.

In agreement with other studies^11^, the mean differences found in the present study between self-reported and measured data were moderate on average (around 1kg/m^2^ for BMI). However, it is important to look beyond differences in means, as moderate differences on average can be accompanied by (i) a large degree of variability between individuals (shown by the SD of the difference between self-reported and measured data)^4^, (ii) a compression of the BMI distribution (shown by lower values at the highest percentiles for self-reported than for measured data)^11^, and (iii) sizeable (downward bias) misclassification of BMI categories based on self-reported data due to such compression (e.g. shifting adults below the BMI cut-off of 30kg/m^2^ for obesity), resulting in an underestimation of obesity prevalence^11^. As reported elsewhere^16^, a large degree of misclassification can occur if a non-trivial number of adults have a moderate difference between self-reported and measured BMI at the margins of broadly defined BMI categories. The positively skewed distribution of BMI increases this effect.

### Prediction equations

Our results were mainly in agreement with previous studies^9 16^: developing prediction equations to correct self-reported height and weight by sociodemographic and health-related variables to more closely approximate measured values of height and weight is feasible. First, in our main analysis, corrected BMI reduced the underestimation of obesity prevalence compared with self-reported BMI^9^, but it remained underestimated (in absolute terms) by 0.8pp for both sexes. As found elsewhere^16^, measured BMI significantly predicted the difference between corrected and measured BMI, indicating that the systematic error in self-reported BMI was not eliminated by the prediction equations. The presence of such residual bias has been identified as a reason for not using equations to predict measured values of height and weight from self-reported values of height and weight^7^. However, the usefulness of prediction equations has been demonstrated by the ability to reduce considerably, although not eliminate, the differences between self-reported and measured anthropometrics across a few, easily gathered sociodemographic and health-related variables^16^, as well as increasing the sensitivity of obesity classification versus self-reported data^8^. Our results also showed that the prediction equations decreased sensitivity in the normal weight category: through erroneously shifting a proportion of normal weight participants to the overweight category, leading to slight overestimation of levels of excess weight. This finding was consistent with previous studies^9 20^, and likely reflects higher levels of reporting accuracy of height and weight among normal weight adults.

Secondly, as elsewhere^8 9 25 35^, our similar results based on full and reduced models, and those of a simpler approach predicting measured values directly from self-reported values and age, confirmed that no single model stood out as the best overall candidate, and that adding variables such as ethnic group and educational status to equations only marginally improved predictive accuracy (shown by similarity in R-squared and estimates of sensitivity). As reported elsewhere^36^, differences between demographic subgroups in the difference between self-reported and measured mean weight may be explained to some extent by differences in measured weight: adjustment for weight in regression models therefore results in attenuation of subgroup differences^37^. It may be reasonably concluded that including additional variables such as educational status and ethnic group does not add enough predictive power to the models to justify the added complexity of including them in prediction equations.

### Strengths and limitations

Pooling data across six years ensured a sample size large enough to compare self-reported and measured height and weight overall and by various sociodemographic and health-related variables, and allowed splitting the data into training and test datasets. Using a regression-based approach we were able to correct for differences in misreporting of height and weight across various subgroups. Unlike other studies^6^, there was no time lapse between the collection of self-reported and measured height and weight, and consistent methodology was used in each survey. We used two approaches to develop prediction equations to enable researchers to evaluate for themselves whether either approach, and if so which, best suits their data and goals.

A study limitation is the sizeable number of participants excluded from our analyses due to missing data on height and weight. Our findings could be biased if complete cases were systematically different from those with missing data (e.g. if those who refused to be measured were more likely than those who did not refuse to underestimate their weight due at least partly to being heavier), and such bias could result in prediction equations that are inaccurate^20^. To partially evaluate this bias we compared obesity prevalence based on self-reported BMI among those with and without measured BMI^25^. Obesity prevalence via self-reported BMI was higher for those without measured BMI (22% men; 25% women) than for those with measured BMI (18% men; 19% women), indicating that heavier participants were less likely to agree to direct measurement^25^. Furthermore, as in other studies, in developing the prediction equations we excluded a small but non-trivial number of participants with a large observed difference between self-reported and measured data: this exclusion may have limited the generalisability of our analyses to some extent. Such cases would be impossible to identify and exclude in surveys that collect self-report but not measured data^13^.

Other limitations include potentially relevant variables that we could not include in regression models due to not being available in all HSE years (e.g. physical activity; perceptions of weight). We also decided *a priori* to use age as a categorical rather than continuous variable in our main analysis. As only categorical age is now provided on publicly available HSE datasets (to preserve anonymity of participants), our approach enables researchers to easily replicate our results and revise/update equations accordingly. (Continuous age was used in our sensitivity analysis to replicate the approach used to correct self-reported height and weight in the ALS). Finally, although we showed no linear trend in misreporting, the prediction equations we have developed based on 2011-16 data might not be entirely applicable to more contemporaneous data. This might be the case if obesity prevalence has greatly increased or decreased, such that the composition of this group changed, and/or the social desirability of having a normal BMI increased or decreased. Changes in accuracy of home scales, or in the up-to-date knowledge of one ‘s own height and weight (e.g. if health-workers began to routinely measure height as part of BMI assessment, and relay that information to patients) could also affect the applicability of these equations to more recent data. Likewise, any potential increase in misreporting of weight associated with weight gain during the Covid-19 pandemic (e.g. due to fewer opportunities for outdoor physical activity) is not taken into account by the equations developed herein.

Our findings must also be interpreted with caution. It is likely that HSE 2011-16 participants might have anticipated that interviewers would take direct measurements of height and weight, resulting in more “truthful” reporting compared, for example, with a telephone interview where participants would not anticipate being measured^10^. Previous studies have shown that misreporting of height (except for older adults) and weight was smaller for in-house interviews compared with telephone interviews^35^. More “truthful” reporting is associated with an underestimation of the differences between self-reported and measured height and weight^3^. Applying the prediction equations developed in the present study on surveys which collect height and weight data by telephone interviews or mailed questionnaires would likely underestimate obesity prevalence to a greater extent than shown herein. Finally, as cautioned elsewhere^9 11 20^, prediction equations are specific to time, place, target population, and methods of data collection. We do not assume that these equations developed using HSE data are applicable to non-HSE samples with different sociodemographic, health and self-reported anthropometric profiles.

## CONCLUSIONS

The prediction equations developed in the present study improved the sensitivity of self-reported obesity and took into account the variations in potential misreporting of height and weight by sociodemographic and health-related variables. Including additional sociodemographic variables does not add enough predictive power to justify the added complexity of including them in prediction equations. Potentially these equations could be used to adjust for errors in self-reported BMI, however important caveats to their use need to be considered.

## Data Availability

The HSE datasets generated and analysed during the current study (age banding for participants) are available via the UK Data Service (UKDS: https://ukdataservice.ac.uk/), subject to their end user license agreement.

## Availability of data and materials

### Abbreviations

BMI: body mass index
HSE: Health Survey for England;
PP: percentage points;
SD: standard deviation

## Acknowledgements

The authors thank the interviewers and nurses, the participants in the Health Survey for England series, and colleagues at NatCen Social Research. The authors also thank NHS Digital. NHS Digital is the trading name of the Health and Social Care Information Centre.

## Contributions

SS conceptualised the study and was responsible for conducting the analyses, interpreting the results and drafting the manuscript. SS, LNF, AM and JSM critically revised the manuscript. All authors have read and approved the manuscript.

## Ethics approval

The procedures used in the HSE to obtain informed consent from survey participants are very closely scrutinised by a NHS ethics committee each year. Approval was obtained from the following Research Ethics Committees (REC): HSE 2011 and 2012: Oxford A REC: 10/H0604/56; HSE 2013 and 2014: Oxford A REC: 12/sc/0317; HSE 2015: West London NRES Committee: 14/LO/0862; HSE 2016: Nottingham REC: 15/EE/0299. This study is a secondary analysis of previously collected data and so additional ethical approval was not required.

