## Supplementary Data for "Does the use of prediction equations to correct self-reported height and weight improve obesity prevalance estimates? A pooled cross-sectional analysis of Health Survey for England data" (from another version of this preprint: version 1, 2022-01-30)

**List of Tables**

| **Table S1** | Differences in means for self-reported and measured height, weight and BMI by survey year and sex | p.2 |
| --- | --- | --- |
| **Table S2** | Means (95% CI) of self-reported, measured and difference of height, weight and BMI by categories of measured BMI | p.3 |
| **Table S3** | Selected percentiles and interquartile range (IQR) of height, weight and BMI by sex | p.4 |
| **Table S4** | Linear regression results for discrepancy between self-reported and measured height (full and reduced models) | pp.5-7 |
| **Table S5** | Linear regression results for discrepancy between self-reported and measured weight (full and reduced models) | pp.8-9 |
| **Table S6** | Regression of discrepancy between corrected and measured BMI on measured BMI by sex | p.10 |
| **Table S7** | Regression of measured height on self-reported height and age by sex | p.11 |
| **Table S8** | Regression of measured height on self-reported height and age by sex | p.12 |
| **Table S9** | Cross-tabulation of measured and corrected BMI categories by sex | p.13 |

**List of Figures**

| **Figure S1** | Bland-Altman plot of the difference between self-reported and measured BMI vs. the average of these two measures of BMI | p.14 |
| --- | --- | --- |
| **Figure S2** | Distributions of self-reported and measured BMI | p.15 |
| **Figure S3** | Bland-Altman plot of the difference between corrected and measured BMI vs. the average of these two measures of BMI | p.16 |
| **Figure S4** | Self-reported, measured and corrected height and weight by age-group and sex | p.17 |

**Table S1	Differences in means for self-reported and measured height, weight and BMI by survey year and sex**

|  | **Difference (95% CI)^a^** | | | | | |
| --- | --- | --- | --- | --- | --- | --- |
|  | **2011** | **2012** | **2013** | **2014** | **2015** | **2016** |
| **Men** |  |  |  |  |  |  |
| Height: mean (cm) | 1.6 (1.5 to 1.7) | 1.6 (1.4 to 1.7) | 1.5 (1.4 to 1.6) | 1.6 (1.5 to 1.7) | 1.7 (1.6 to 1.9) | 1.5 (1.4 to 1.6) |
| Weight: mean (kg) | -1.5 (-1.6 to -1.3) | -1.6 (-1.8 to -1.4) | -1.5 (-1.6 to -1.3) | -1.3 (-1.5 to -1.1) | -1.8 (-2.0 to -1.6) | -1.5 (-1.7 to -1.3) |
| BMI: mean (kg/m^2^) | -1.0 (-1.0 to -0.9) | -1.0 (-1.1 to -0.9) | -0.9 (-1.0 to -0.9) | -0.9 (-1.0 to -0.8) | -1.1 (-1.2 to -1.0) | -1.0 (-1.0 to -0.9) |
| *Overweight incl. obese (%)^b^* | *-7.8pp (-9.1 to -6.5)* | *-8.6pp (-9.9 to -7.3)* | *-8.6pp (-9.9 to -7.3)* | *-7.4pp (-8.8 to -6.1)* | *-8.3pp (-9.6 to -7.0)* | *-7.3pp (-8.6 to -6.0)* |
| *Obese (%)^c^* | *-6.8pp (-7.9 to -5.7)* | *-6.4pp (-7.5 to -5.4)* | *-6.6pp (-7.6 to -5.6)* | *-6.1pp (-7.1 to -5.0)* | *-7.7pp (-8.8 to -6.6)* | *-7.2pp (-8.4 to -6.1)* |
| **Women** |  |  |  |  |  |  |
| Height: mean (cm) | 1.0 (0.9 to 1.1) | 1.0 (0.9 to 1.1) | 0.9 (0.8 to 1.0) | 1.0 (0.9 to 1.1) | 1.1 (1.0 to 1.2) | 1.0 (0.9 to 1.1) |
| Weight: mean (kg) | -2.1 (-2.2 to -1.9) | -2.1 (-2.2 to -1.9) | -2.0 (-2.2 to -1.9) | -2.0 (-2.2 to -1.8) | -2.2 (-2.4 to -2.0) | -2.0 (-2.2 to -1.8) |
| BMI: mean (kg/m^2^) | -1.1 (-1.2 to -1.1) | -1.1 (-1.2 to -1.0) | -1.1 (-1.1 to -1.0) | -1.1 (-1.2 to -1.0) | -1.2 (-1.2 to -1.1) | -1.1 (-1.2 to -1.0) |
| *Overweight incl. obese (%)^b^* | *-9.6pp (-10.7 to -8.5)* | *-8.4pp (-9.5 to -7.3)* | *-10.0pp (-11.1 to -8.9)* | *-8.8pp (-10.0 to -7.7)* | *-8.0pp (-9.1 to -7.0)* | *-8.7pp (-9.9 to -7.5)* |
| *Obese (%)^c^* | *-5.7pp (-6.6 to -4.8)* | *-5.7pp (-6.6 to -4.8)* | *-5.6pp (-6.5 to -4.7)* | *-6.9pp (-7.8 to -5.9)* | *-6.5pp (-7.4 to -5.6)* | *-5.8pp (-6.7 to -4.8)* |

BMI: body mass index; CI: confidence interval; PP: percentage points. Participants with valid self-reported and measured height and weight (n=38,942).

**^a^** Self-reported minus measured.

^b^ Overweight including obese: BMI ≥25.0kg/m^2^.

^c^ Obese: BMI ≥30.0kg/m^2^.

**Table S2	Means (95% CI) of self-reported, measured and difference of height, weight and BMI by categories of measured BMI**

|  | **Measured BMI categories** | | | | |
| --- | --- | --- | --- | --- | --- |
|  | **Underweight** | **Normal** | **Overweight but not obese** | **Obese I & II** | **Obese III** |
| **Men** |  |  |  |  |  |
| Height_SR_: mean (cm) | 176.4 (175.1 to 177.7) | 177.6 (177.4 to 177.8) | 176.9 (176.7 to 177.1) | 176.9 (176.6 to 177.1) | 177.2 (176.3 to 178.2) |
| Height_M_: mean (cm) | 175.2 (174.1 to 176.4) | 176.4 (176.2 to 176.7) | 175.3 (175.1 to 175.4) | 174.9 (174.7 to 175.1) | 174.8 (173.9 to 175.8) |
| *Height_DIFF_: mean (cm)* | *1.2 (0.6 to 1.8)* | *1.2 (1.1 to 1.3)* | *1.7 (1.6 to 1.7)* | *2.0 (1.9 to 2.1)* | *2.4 (2.0 to 2.7)* |
| Weight_SR_: mean (kg) | 55.7 (54.7 to 56.7) | 70.8 (70.5 to 71.0) | 82.5 (82.3 to 82.7) | 98.4 (98.1 to 98.8) | 127.2 (125.3 to 129.2) |
| Weight_M_: mean (kg) | 53.8 (53.0 to 54.6) | 70.9 (70.6 to 71.1) | 84.2 (84.0 to 84.4) | 101.5 (101.1 to 101.9) | 134.0 (131.9 to 136.0) |
| *Weight_DIFF_: mean (kg)* | *1.9 (1.3 to 2.5)* | *-0.1 (-0.2 to 0.0)* | *-1.6 (-1.7 to -1.5)* | *-3.0 (-3.2 to -2.9)* | *-6.7 (-7.9 to -5.5)* |
| BMI_SR_: mean (kg/m^2^) | 17.9 (17.6 to 18.1) | 22.4 (22.4 to 22.5) | 26.3 (26.3 to 26.4) | 31.4 (31.3 to 31.5) | 40.5 (39.9 to 41.1) |
| BMI_M_: mean (kg/m^2^) | 17.5 (17.4 to 17.6) | 22.7 (22.7 to 22.8) | 27.4 (27.3 to 27.4) | 33.1 (33.0 to 33.2) | 43.7 (43.2 to 44.3) |
| *BMI_DIFF_: mean (kg/m^2^)* | *0.4 (0.2 to 0.7)* | *-0.3 (-0.4 to -0.3)* | *-1.0 (-1.1 to -1.0)* | *-1.7 (-1.8 to -1.6)* | *-3.2 (-3.7 to -2.8)* |
| **Women** |  |  |  |  |  |
| Height_SR_: mean (cm) | 163.1 (162.2 to 163.9) | 163.5 (163.4 to 163.7) | 162.7 (162.5 to 162.9) | 162.0 (161.7 to 162.2) | 162.5 (162.0 to 163.0) |
| Height_M_: mean (cm) | 162.5 (161.7 to 163.4) | 162.8 (162.7 to 163.0) | 161.5 (161.4 to 161.7) | 160.6 (160.4 to 160.8) | 161.2 (160.6 to 161.7) |
| *Height_DIFF_: mean (cm)* | *0.5 (0.1 to 0.9)* | *0.7 (0.6 to 0.8)* | *1.2 (1.1 to 1.3)* | *1.3 (1.2 to 1.4)* | *1.3 (1.1 to 1.6)* |
| Weight_SR_: mean (cm) | 47.4 (46.8 to 48.0) | 58.3 (58.2 to 58.5) | 69.1 (68.9 to 69.3) | 83.2 (82.9 to 83.5) | 107.7 (106.5 to 108.9) |
| Weight_M_: mean (cm) | 46.5 (46.0 to 47.1) | 59.3 (59.2 to 59.5) | 71.3 (71.1 to 71.5) | 86.8 (86.5 to 87.1) | 113.8 (112.7 to 114.8) |
| *Weight_DIFF_: mean (cm)* | *0.9 (0.5 to 1.2)* | *-1.0 (-1.1 to -1.0)* | *-2.2 (-2.3 to -2.1)* | *-3.6 (-3.8 to -3.4)* | *-6.1 (-6.8 to -5.3)* |
| BMI_SR_: mean (kg/m^2^) | 17.8 (17.7 to 18.0) | 21.8 (21.7 to 21.8) | 26.1 (26.0 to 26.1) | 31.7 (31.6 to 31.8) | 40.8 (40.4 to 41.1) |
| BMI_M_: mean (kg/m^2^) | 17.6 (17.5 to 17.6) | 22.3 (22.3 to 22.4) | 27.3 (27.2 to 27.3) | 33.6 (33.5 to 33.6) | 43.7 (43.4 to 44.0) |
| *BMI_DIFF_: mean (kg/m^2^)* | *0.2 (0.1 to 0.4)* | *-0.6 (-0.6 to -0.5)* | *-1.2 (-1.2 to -1.2)* | *-1.9 (-1.9 to -1.8)* | *-3.0 (-3.3 to -2.7)* |

SR: self-reported; M: measured; DIFF: difference (SR – M). Participants with valid self-reported and measured height and weight (n=38,942).

**Table S3	Selected percentiles and interquartile range (IQR) of height, weight and BMI by sex**

|  | 5^th^ | 10^th^ | 25^th^ | 50^th^ | 75^th^ | 90^th^ | 95^th^ | IQR |
| --- | --- | --- | --- | --- | --- | --- | --- | --- |
|  | **Height (cm)** | | | | | | | |
| **Men** |  |  |  |  |  |  |  |  |
| Measured | 163.5 | 166.2 | 170.7 | 175.5 | 180.4 | 184.8 | 187.5 | 9.7 |
| Self-report | 165.1 | 167.6 | 172.7 | 177.8 | 182.9 | 186.0 | 188.0 | 10.2 |
| **Women** |  |  |  |  |  |  |  |  |
| Measured | 150.5 | 153.0 | 157.3 | 162.0 | 166.5 | 170.6 | 173.0 | 9.2 |
| Self-report | 152.4 | 154.9 | 157.5 | 162.6 | 167.6 | 172.7 | 175.0 | 10.1 |
|  | **Weight (kg)** | | | | | | | |
| **Men** |  |  |  |  |  |  |  |  |
| Measured | 61.3 | 65.9 | 73.3 | 82.5 | 93.5 | 105.4 | 113.8 | 20.2 |
| Self-report | 60.8 | 65.0 | 72.2 | 81.3 | 91.7 | 102.0 | 111.2 | 19.5 |
| **Women** |  |  |  |  |  |  |  |  |
| Measured | 50.2 | 53.6 | 59.6 | 67.9 | 78.5 | 90.9 | 99.9 | 18.9 |
| Self-report | 50.0 | 52.2 | 58.0 | 66.0 | 76.3 | 88.0 | 95.3 | 18.3 |
|  | **BMI (kg/m^2^)** | | | | | | | |
| **Men** |  |  |  |  |  |  |  |  |
| Measured | 20.4 | 21.7 | 24.1 | 26.8 | 30.0 | 33.6 | 36.3 | 6.0 |
| Self-report | 20.0 | 21.3 | 23.4 | 25.9 | 28.8 | 32.2 | 34.6 | 5.3 |
| **Women** |  |  |  |  |  |  |  |  |
| Measured | 19.6 | 20.6 | 22.8 | 25.8 | 29.9 | 34.7 | 37.9 | 7.1 |
| Self-report | 19.1 | 20.0 | 22.0 | 24.8 | 28.5 | 32.9 | 36.2 | 6.5 |

BMI: body mass index. IQR: inter-quartile range: [75^th^ minus 25^th^ percentile]. Participants with valid self-reported and measured height and weight (n=38,942).

**Table S4	Linear regression results for difference between self-reported and measured height (full and reduced models)**

| **Predictors of difference** | **Full model** | | |  | **Reduced model** | | |
| --- | --- | --- | --- | --- | --- | --- | --- |
|  | **β** | **95% CI** | ***P*** |  | **β** | **95% CI** | ***P*** |
|  | *Men (n=12,376)* | | | | | | |
| Intercept | -100.572 | -123.955 to -77.190 | <0.001 |  | -92.647 | -115.865 to -69.428 | <0.001 |
| Height_(self-report)_ | **1.040** | **0.777 to 1.303** | **<0.001** |  | **0.963** | **0.701 to 1.224** | **<0.001** |
| Height^2^_(self-report)_ | **-0.003** | **-0.003 to -0.002** | **<0.001** |  | **-0.002** | **-0.003 to -0.002** | **<0.001** |
| **Age-group (p<0.001):** | | | |  |  |  |  |
| 16-17y | REF | - | - |  | REF | - | - |
| 18-19y | 0.460 | -0.028 to 0.948 | 0.065 |  | 0.403 | -0.085 to 0.891 | 0.105 |
| 20-24y | 0.119 | -0.310 to 0.548 | 0.588 |  | 0.035 | -0.395 to 0.464 | 0.874 |
| 25-29y | 0.055 | -0.355 to 0.465 | 0.793 |  | -0.056 | -0.462 to 0.350 | 0.787 |
| 30-34y | -0.247 | -0.645 to 0.151 | 0.224 |  | -0.358 | -0.753 to 0.037 | 0.076 |
| 35-39y | -0.027 | -0.428 to 0.374 | 0.895 |  | -0.135 | -0.533 to 0.262 | 0.504 |
| 40-44y | 0.369 | -0.024 to 0.761 | 0.065 |  | 0.253 | -0.137 to 0.642 | 0.204 |
| 45-49y | 0.392 | -0.006 to 0.790 | 0.054 |  | 0.312 | -0.083 to 0.707 | 0.121 |
| 50-54y | **0.470** | **0.077 to 0.863** | **0.019** |  | 0.389 | 0.000 to 0.779 | 0.050 |
| 55-59y | **0.826** | **0.427 to 1.225** | **<0.001** |  | **0.734** | **0.340 to 1.129** | **<0.001** |
| 60-64y | **1.103** | **0.707 to 1.499** | **<0.001** |  | **1.001** | **0.611 to 1.391** | **<0.001** |
| 65-69y | **1.568** | **1.173 to 1.963** | **<0.001** |  | **1.463** | **1.073 to 1.853** | **<0.001** |
| 70-74y | **2.153** | **1.734 to 2.572** | **<0.001** |  | **2.089** | **1.678 to 2.500** | **<0.001** |
| 75-79y | **2.516** | **2.091 to 2.942** | **<0.001** |  | **2.452** | **2.032 to 2.872** | **<0.001** |
| 80-84y | **2.926** | **2.443 to 3.410** | **<0.001** |  | **2.850** | **2.372 to 3.329** | **<0.001** |
| 85y+ | **4.103** | **3.548 to 4.657** | **<0.001** |  | **4.026** | **3.479 to 4.573** | **<0.001** |
| **Region (p=0.033):** | | | |  |  |  |  |
| North East | **0.316** | **0.098 to 0.533** | **0.004** |  | - | - | - |
| North West | **0.199** | **0.014 to 0.384** | **0.035** |  | - | - | - |
| Yorkshire | 0.196 | -0.009 to 0.401 | 0.061 |  | - | - | - |
| East Midlands | 0.023 | -0.173 to 0.218 | 0.821 |  | - | - | - |
| West Midlands | **0.296** | **0.089 to 0.503** | **0.005** |  | - | - | - |
| East of England | 0.192 | -0.002 to 0.386 | 0.053 |  | - | - | - |
| London | 0.122 | -0.066 to 0.310 | 0.205 |  | - | - | - |
| South East | REF | - | - |  | - | - | - |
| South West | 0.070 | -0.121 to 0.262 | 0.472 |  | - | - | - |
| **Education (p<0.001):** | | | |  |  |  |  |
| Degree | REF | - | - |  | - | - | - |
| Below degree | **0.143** | **0.019 to 0.266** | **0.023** |  | - | - | - |
| O level/other | **0.276** | **0.143 to 0.408** | **<0.001** |  | - | - | - |
| None | **0.244** | **0.074 to 0.414** | **0.005** |  | - | - | - |
| Missing | 0.603 | -0.738 to 1.943 | 0.378 |  | - | - | - |
| **Ethnic group (p<0.001):** | | | |  |  |  |  |
| White | REF | - | - |  | - | - | - |
| Black | -0.299 | -0.656 to 0.057 | 0.100 |  | - | - | - |
| Asian | **0.605** | **0.361 to 0.850** | **<0.001** |  | - | - | - |
| Mixed | 0.385 | -0.049 to 0.819 | 0.082 |  | - | - | - |
| Other | 0.153 | -0.494 to 0.800 | 0.643 |  | - | - | - |
| Missing | -0.264 | -2.519 to 1.991 | 0.819 |  | - | - | - |
| **General health (p<0.001):** | | | |  |  |  |  |
| Very good/good | REF | - | - |  | - | - | - |
| Fair | 0.126 | -0.011 to 0.264 | 0.072 |  | - | - | - |
| Very bad/bad | **0.460** | **0.239 to 0.681** | **<0.001** |  | - | - | - |
| **R^2^ (%)** | **16.1** |  |  |  | **15.2** |  |  |

**TABLE S4…..continued**

| **Predictors of difference** | **Full model** | | |  | **Reduced model** | | |
| --- | --- | --- | --- | --- | --- | --- | --- |
|  | **β** | **95% CI** | ***P*** |  | **β** | **95% CI** | ***P*** |
|  | *Women (n=14,659)* | | | | | | |
| Intercept | -21.809 | -23.070 to -20.548 | <0.001 |  | -18.150 | -19.355 to -16.944 | <0.001 |
| Height_(self-report)_ | 0.135 | 0.128 to 0.142 | <0.001 |  | 0.117 | 0.110 to 0.124 | <0.001 |
| **Age (p<0.001):** |  |  |  |  |  |  |  |
| 16-17y | REF | - | - |  | REF | - | - |
| 18-19y | -0.221 | -0.688 to 0.245 | 0.352 |  | -0.329 | -0.801 to 0.144 | 0.173 |
| 20-24y | **-0.515** | **-0.903 to -0.127** | **0.009** |  | **-0.644** | **-1.028 to -0.260** | **0.001** |
| 25-29y | **-0.563** | **-0.927 to -0.200** | **0.002** |  | **-0.712** | **-1.075 to -0.350** | **<0.001** |
| 30-34y | -0.310 | -0.679 to 0.059 | 0.100 |  | **-0.388** | **-0.755 to -0.020** | **0.039** |
| 35-39y | -0.108 | -0.471 to 0.255 | 0.560 |  | -0.237 | -0.600 to 0.126 | 0.201 |
| 40-44y | -0.129 | -0.487 to 0.229 | 0.481 |  | -0.280 | -0.637 to 0.077 | 0.124 |
| 45-49y | -0.114 | -0.473 to 0.246 | 0.536 |  | -0.318 | -0.675 to 0.040 | 0.082 |
| 50-54y | -0.147 | -0.505 to 0.212 | 0.423 |  | -0.366 | -0.725 to -0.008 | 0.045 |
| 55-59y | 0.268 | -0.092 to 0.627 | 0.144 |  | 0.008 | -0.352 to 0.367 | 0.967 |
| 60-64y | **0.676** | **0.314 to 1.037** | **<0.001** |  | **0.449** | **0.089 to 0.809** | **0.015** |
| 65-69y | **1.057** | **0.686 to 1.428** | **<0.001** |  | **0.848** | **0.479 to 1.218** | **<0.001** |
| 70-74y | **1.607** | **1.225 to 1.989** | **<0.001** |  | **1.398** | **1.019 to 1.776** | **<0.001** |
| 75-79y | **2.349** | **1.946 to 2.751** | **<0.001** |  | **2.171** | **1.774 to 2.569** | **<0.001** |
| 80-84y | **3.527** | **3.068 to 3.986** | **<0.001** |  | **3.362** | **2.904 to 3.820** | **<0.001** |
| 85+y | **4.370** | **3.823 to 4.917** | **<0.001** |  | **4.179** | **3.632 to 4.727** | **<0.001** |
| **Region (p=0.023)** | | |  |  |  |  |  |
| North East | 0.093 | -0.088 to 0.274 | 0.315 |  | - | - | - |
| North West | **0.261** | **0.099 to 0.423** | **0.002** |  | - | - | - |
| Yorkshire | 0.158 | -0.021 to 0.337 | 0.084 |  | - | - | - |
| East Midlands | 0.039 | -0.155 to 0.233 | 0.694 |  | - | - | - |
| West Midlands | 0.131 | -0.051 to 0.314 | 0.158 |  | - | - | - |
| East | -0.048 | -0.204 to 0.108 | 0.546 |  | - | - | - |
| London | 0.046 | -0.123 to 0.215 | 0.594 |  | - | - | - |
| South East | REF | - | - |  | - | - | - |
| South West | 0.015 | -0.144 to 0.174 | 0.854 |  | - | - | - |
| **Education (p<0.001):** | | |  |  |  |  |  |
| Degree | REF | - | - |  | - | - | - |
| Below degree | 0.097 | -0.012 to 0.206 | 0.081 |  | - | - | - |
| O level/other | **0.186** | **0.073 to 0.299** | **0.001** |  | - | - | - |
| None | **0.519** | **0.367 to 0.671** | **<0.001** |  | - | - | - |
| Missing | 0.368 | -0.883 to 1.619 | 0.564 |  | - | - | - |
| **Index of Multiple Deprivation (p=0.007):** | | |  |  |  |  |  |
| Least deprived | REF | - | - |  | - | - | - |
| 2 | 0.050 | -0.071 to 0.172 | 0.414 |  | - | - | - |
| 3 | 0.100 | -0.029 to 0.228 | 0.128 |  | - | - | - |
| 4 | **0.252** | **0.114 to 0.390** | **<0.001** |  | - | - | - |
| Most deprived | 0.154 | -0.001 to 0.308 | 0.052 |  | - | - | - |
| **Ethnic group (p<0.001):** | | | |  |  |  |  |
| White | REF | - | - |  | - | - | - |
| Black | **0.529** | **0.179 to 0.878** | **0.003** |  | - | - | - |
| Asian | **1.785** | **1.572 to 1.997** | **<0.001** |  | - | - | - |
| Mixed | **0.500** | **0.044 to 0.957** | **0.032** |  | - | - | - |
| Other | 1.696 | 1.228 to 2.163 | <0.001 |  | - | - | - |
| Missing | -0.066 | -1.654 to 1.523 | 0.935 |  | - | - | - |
| R^2^ (%) | **22.3** |  |  |  | **18.6** |  |  |

*Notes:* REF: reference group. Dependent variable: difference between self-reported and measured height. Height is in centimetres. Parameters estimated using split-sample A.

*_________________________________________________________________________*

*Prediction equations:* using the same set of independent variables in a linear regression model with measured values of height as the dependent variable results in the regression parameters below. These can be used to obtain an estimate of the corrected height measurements for participants.

**Adjusted height: full model** (men) = 100.572 – 0.040×(height_self-reported_) + 0.003×(height^2^_self-reported_) – 0.460×(age 18-19y) - 0.119×(age 20-24y) - 0.055×(age 25-29y) + 0.247×(age 30-34y) + 0.027×(age 35-39y) – 0.369×(age 40-44y) – 0.392×(age 45-49y) – 0.470×(age 50-54y) – 0.826×(age 55-59y) – 1.103×(age 60-64y) – 1.568×(age 65-69y) – 2.153×(age 70-74y) - 2.516×(age 75-79y) - 2.926×(age 80-84y) – 4.103×(age 85y+) – 0.316×(North East) - 0.199×(North West) - 0.196×(Yorkshire & The Humber) - 0.023×(East Midlands) - 0.296×(West Midlands) - 0.192×(East of England) - 0.122×(London) - 0.070×(South West) – 0.143×(below degree) – 0.276×(O level/GCSE) – 0.244×(none) – 0.603×(missing education data) + 0.299×(Black) - 0.605×(Asian) - 0.385×(Mixed) - 0.153×(Other ethnic group) + 0.264×(missing ethnic data) -0.126×(fair general health) – 0.460×(bad/very bad general health) (R^2^=88.6%)

**Adjusted height: reduced model** (men) = 92.647 + 0.037×(height_self-reported_) + 0.002×(height^2^_self-reported_) – 0.403×(age 18-19y) - 0.035×(age 20-24y) + 0.056×(age 25-29y) + 0.358×(age 30-34y) + 0.135×(age 35-39y) - 0.253×(age 40-44y) – 0.312×(age 45-49y) – 0.389×(age 50-54y) – 0.734×(age 55-59y) – 1.001×(age 60-64y) – 1.463×(age 65-69y) – 2.089×(age 70-74y) - 2.452×(age 75-79y) - 2.850×(age 80-84y) – 4.026×(age 85y+) (R^2^=88.5%)

**Adjusted height: full model** (women) = 21.809 + 0.865×(height_self-reported_) + 0.221×(age 18-19y) + 0.515×(age 20-24y) + 0.563×(age 25-29y) + 0.310×(age 30-34y) + 0.108×(age 35-39y) + 0.129×(age 40-44y) + 0.114×(age 45-49y) + 0.147×(age 50-54y) – 0.268×(age 55-59y) – 0.676×(age 60-64y) – 1.057×(age 65-69y) – 1.607×(age 70-74y) - 2.349×(age 75-79y) - 3.527×(age 80-84y) – 4.370×(age 85y+) - 0.093×(North East) - 0.261×(North West) - 0.158×(Yorkshire & The Humber) - 0.039×(East Midlands) - 0.131×(West Midlands) + 0.048×(East of England) - 0.046×(London) - 0.015×(South West) – 0.097×(below degree) – 0.186×(O level/GCSE) – 0.519×(none) - 0.368×(missing education data) - 0.050×(IMD Q2) - 0.100×(IMD Q3) - 0.252×(IMD Q4) - 0.154×(IMD Q5) - 0.529×(Black) - 1.785×(Asian) - 0.500×(Mixed) - 1.696×(Other) + 0.066×(missing ethnic data) (R^2^=87.7%)

**Adjusted height: reduced model** (women) = 18.150 + 0.883×(height_self-reported_) + 0.329×(age 18-19y) + 0.644×(age 20-24y) + 0.712×(age 25-29y) + 0.388×(age 30-34y) + 0.237×(age 35-39y) + 0.280×(age 40-44y) + 0.318×(age 45-49y) + 0.366×(age 50-54y) – 0.008×(age 55-59y) – 0.449×(age 60-64y) – 0.848×(age 65-69y) – 1.398×(age 70-74y) – 2.171×(age 75-79y) - 3.362×(age 80-84y) – 4.179×(age 85y+) (R^2^=87.1%)

*Note:* Height values are in centimetres. Reference groups: age-group (16-17y); Government Office Region (South East); educational status (degree or equivalent); ethnic group (White); smoking status (current cigarette smoker); general health (very good/good).

**Table S5	Linear regression results for difference between self-reported and measured weight (full and reduced models)**

| **Predictors of difference** | **Full model** | | |  | **Reduced model** | | |
| --- | --- | --- | --- | --- | --- | --- | --- |
|  | **β** | **95% CI** | ***P*** |  | **β** | **95% CI** | ***P*** |
|  | *Men (n=12,376)* | | | | | | |
| Intercept | -7.555 | -12.074 to -3.035 | 0.001 |  | -6.931 | -11.371 to -2.491 | 0.002 |
| Weight_(self-report)_ | 0.230 | 0.085 to 0.376 | 0.002 |  | 0.204 | 0.060 to 0.347 | 0.005 |
| Weight^2^_(self-report)_ | -0.003 | -0.004 to -0.001 | 0.001 |  | -0.002 | -0.004 to -0.001 | 0.002 |
| Weight^3^_(self-report)_ | 8.28e-06 | 3.22e06 to 0.000 | 0.001 |  | 7.69e-06 | 2.67e-06 to 0.000 | 0.003 |
| **Age (p<0.001):** |  |  |  |  |  |  |  |
| 16-17y | REF | - | - |  | REF | - | - |
| 18-19y | -0.488 | -1.365 to 0.390 | 0.276 |  | -0.435 | -1.294 to 0.425 | 0.321 |
| 20-24y | 0.450 | -0.260 to 1.159 | 0.215 |  | 0.494 | -0.210 to 1.197 | 0.169 |
| 25-29y | 0.336 | -0.356 to 1.028 | 0.341 |  | 0.400 | -0.277 to 1.078 | 0.247 |
| 30-34y | 0.545 | -0.135 to 1.225 | 0.116 |  | 0.586 | -0.074 to 1.246 | 0.082 |
| 35-39y | 0.231 | -0.432 to 0.894 | 0.495 |  | 0.243 | -0.402 to 0.889 | 0.460 |
| 40-44y | -0.033 | -0.709 to 0.642 | 0.923 |  | -0.012 | -0.673 to 0.649 | 0.971 |
| 45-49y | -0.209 | -0.885 to 0.467 | 0.544 |  | -0.163 | -0.823 to 0.497 | 0.628 |
| 50-54y | -0.289 | -0.954 to 0.375 | 0.393 |  | -0.242 | -0.892 to 0.408 | 0.466 |
| 55-59y | -0.417 | -1.090 to 0.256 | 0.224 |  | -0.377 | -1.035 to 0.280 | 0.261 |
| 60-64y | -0.540 | -1.205 to 0.126 | 0.112 |  | -0.512 | -1.161 to 0.137 | 0.122 |
| 65-69y | -0.324 | -0.994 to 0.345 | 0.342 |  | -0.313 | -0.965 to 0.340 | 0.348 |
| 70-74y | -0.268 | -0.949 to 0.413 | 0.440 |  | -0.237 | -0.896 to 0.422 | 0.481 |
| 75-79y | -0.219 | -0.910 to 0.473 | 0.535 |  | -0.201 | -0.873 to 0.470 | 0.557 |
| 80-84y | 0.042 | -0.690 to 0.773 | 0.911 |  | 0.050 | -0.660 to 0.760 | 0.890 |
| 85+y | -0.303 | -1.107 to 0.501 | 0.460 |  | -0.306 | -1.089 to 0.477 | 0.443 |
| **Education (p=0.010):** | | |  |  |  |  |  |
| Degree | REF | - | - |  | - | - | - |
| Below degree | 0.194 | -0.005 to 0.393 | 0.056 |  | - | - | - |
| O level/other | 0.301 | 0.088 to 0.514 | 0.006 |  | - | - | - |
| None | 0.363 | 0.116 to 0.610 | 0.004 |  | - | - | - |
| Missing | -0.659 | -3.629 to 2.312 | 0.664 |  | - | - | - |
| **Smoking status (p<0.001):** | | |  |  |  |  |  |
| Current | REF | - | - |  | - | - | - |
| Ex-regular | -0.627 | -0.871 to -0.383 | <0.001 |  | - | - | - |
| Never | -0.652 | -0.879 to -0.424 | <0.001 |  | - | - | - |
| Missing | -0.970 | -2.652 to 0.712 | 0.258 |  | - | - | - |
| R^2^ (%) | **2.0** |  |  |  | **1.3** |  |  |

**Table S5….continued**

| **Predictors of difference** | **Full model** | | |  | **Reduced model** | | |
| --- | --- | --- | --- | --- | --- | --- | --- |
|  | **β** | **95% CI** | ***P*** |  | **β** | **95% CI** | ***P*** |
|  | *Women (n=14,659)* | | | | | | |
| Intercept | -2.112 | -5.254 to 1.030 | 0.188 |  | -2.073 | -5.206 to 1.060 | 0.195 |
| **Weight (kg):** |  |  |  |  |  |  |  |
| Weight_(self-report)_ | 0.052 | -0.069 to 0.172 | 0.400 |  | 0.045 | -0.076 to 0.165 | 0.466 |
| Weight^2^_(self-report)_ | -0.001 | -0.003 to 0.000 | 0.076 |  | -0.001 | -0.003 to 0.000 | 0.089 |
| Weight^3^_(self-report)_ | 7.45e-06 | 1.38e-06 to 0.000 | 0.016 |  | 7.30e-06 | 1.23e-06 to 0.000 | 0.018 |
| **Age (p<0.001):** |  |  |  |  |  |  |  |
| 16-17y | REF | - | - |  | REF | - | - |
| 18-19y | 0.821 | 0.186 to 1.456 | 0.011 |  | 0.822 | 0.157 to 1.486 | 0.015 |
| 20-24y | 0.792 | 0.223 to 1.362 | 0.006 |  | 0.834 | 0.224 to 1.444 | 0.007 |
| 25-29y | 1.082 | 0.547 to 1.616 | <0.001 |  | 1.127 | 0.554 to 1.700 | <0.001 |
| 30-34y | 0.939 | 0.406 to 1.473 | 0.001 |  | 0.962 | 0.389 to 1.536 | 0.001 |
| 35-39y | 0.867 | 0.344 to 1.390 | 0.001 |  | 0.891 | 0.327 to 1.455 | 0.002 |
| 40-44y | 0.825 | 0.299 to 1.350 | 0.002 |  | 0.852 | 0.286 to 1.417 | 0.003 |
| 45-49y | 0.564 | 0.036 to 1.092 | 0.036 |  | 0.595 | 0.026 to 1.163 | 0.040 |
| 50-54y | 0.551 | 0.025 to 1.077 | 0.040 |  | 0.584 | 0.018 to 1.151 | 0.043 |
| 55-59y | 0.685 | 0.151 to 1.219 | 0.012 |  | 0.720 | 0.143 to 1.297 | 0.014 |
| 60-64y | 0.651 | 0.121 to 1.180 | 0.016 |  | 0.676 | 0.107 to 1.245 | 0.020 |
| 65-69y | 0.592 | 0.071 to 1.113 | 0.026 |  | 0.612 | 0.050 to 1.173 | 0.033 |
| 70-74y | 0.953 | 0.419 to 1.487 | <0.001 |  | 0.961 | 0.386 to 1.535 | 0.001 |
| 75-79y | 0.562 | 0.004 to 1.120 | 0.049 |  | 0.565 | -0.033 to 1.162 | 0.064 |
| 80-84y | 0.763 | 0.155 to 1.371 | 0.014 |  | 0.768 | 0.124 to 1.413 | 0.019 |
| 85+y | 1.378 | 0.694 to 2.062 | <0.001 |  | 1.377 | 0.664 to 2.091 | <0.001 |
| **Smoking status (p=0.034):** | | |  |  |  |  |  |
| Current | REF | - | - |  | - | - | - |
| Ex-regular | -0.255 | -0.447 to -0.063 | 0.009 |  | - | - | - |
| Never | -0.178 | -0.356 to 0.000 | 0.050 |  | - | - | - |
| Missing | -0.359 | -2.377 to 1.659 | 0.727 |  | - | - | - |
| **Ethnic group (p=0.009):** | | | |  |  |  |  |
| White | REF | - | - |  | - | - | - |
| Black | -0.871 | -1.380 to -0.361 | 0.001 |  | - | - | - |
| Asian | 0.011 | -0.246 to 0.269 | 0.932 |  | - | - | - |
| Mixed | 0.244 | -0.366 to 0.854 | 0.433 |  | - | - | - |
| Other | 0.371 | -0.211 to 0.953 | 0.212 |  | - | - | - |
| Missing | -0.716 | -2.643 to 1.212 | 0.467 |  | - | - | - |
| R^2^ | **1.9** |  |  |  | **1.7** |  |  |

*Notes:* REF: reference group. Weight values are in kilogrammes. Dependent variable: difference between self-reported and measured weight. Parameters estimated using split-sample A.

*_________________________________________________________________________*

*Prediction equations:* using the same set of independent variables in a linear regression model with measured values of weight as the dependent variable results in the regression parameters below. These can be used to obtain an estimate of the corrected weight measurements for participants.

**Adjusted weight:** **full model** (men) = 7.555 + 0.770×(weight_self-reported_) + 0.003×(weight^2^_self-reported_) – 8.28e-06×(weight^3^_self-reported_) + 0.488×(age 18-19y) – 0.450×(age 20-24y) – 0.336×(age 25-29y) - 0.545×(age 30-34y) - 0.231×(age 35-39y) + 0.033×(age 40-44y) + 0.209×(age 45-49y) + 0.289×(age 50-54y) + 0.417×(age 55-59y) + 0.540×(age 60-64y) + 0.324×(age 65-69y) + 0.268×(age 70-74y) + 0.219×(age 75-79y) - 0.042×(age 80-84y) + 0.303×(age 85y+) - 0.194×(below degree) – 0.301×(O level/GCSE) - 0.363×(none) + 0.659×(missing education data) + 0.627×(ex-regular smoker) + 0.652×(never regular smoker) + 0.970×(missing smoking data) (R^2^=94.3%)

**Adjusted weight: reduced model** (men) = 6.931 + 0.796×(weight_self-reported_) + 0.002×(weight^2^_self-reported_) -7.69e-06×(weight^3^_self-reported_) + 0.435×(age 18-19y) – 0.494×(age 20-24y) – 0.400×(age 25-29y) - 0.586×(age 30-34y) - 0.243×(age 35-39y) + 0.012×(age 40-44y) + 0.163×(age 45-49y) + 0.242×(age 50-54y) + 0.377×(age 55-59y) + 0.512×(age 60-64y) + 0.313×(age 65-69y) + 0.237×(age 70-74y) + 0.201×(age 75-79y) - 0.050×(age 80-84y) + 0.306×(age 85y+) (R^2^=94.3%)

**Adjusted weight: full model** (women) 2.112 + 0.948×(weight_self-reported_) + 0.001×(weight^2^_self-reported_) -7.45e-06×(weight^3^_self-reported_) - 0.821×(age 18-19y) - 0.792×(age 20-24y) - 1.082×(age 25-29y) - 0.939×(age 30-34y) - 0.867×(age 35-39y) - 0.825×(age 40-44y) – 0.564×(age 45-49y) - 0.551×(age 50-54y) – 0.685×(age 55-59y) – 0.651×(age 60-64y) – 0.592×(age 65-69y) – 0.953×(age 70-74y) - 0.562×(age 75-79y) - 0.763×(age 80-84y) – 1.378×(age 85y+) + 0.255×(ex-regular smoker) + 0.178×(never regular smoker) + 0.359×(missing smoking data) + 0.871×(Black) - 0.011×(Asian) - 0.244×(Mixed) - 0.371×(Other) + 0.716×(missing ethnic data) (R^2^=95.1%)

**Adjusted weight: reduced model** (women) = 2.073 + 0.955(weight_self-reported_) + 0.001(weight^2^_self-reported_) -7.30e-06(weight^3^_self-reported_) - 0.822(age 18-19y) - 0.834(age 20-24y) - 1.127(age 25-29y) - 0.962(age 30-34y) - 0.891(age 35-39y) - 0.852(age 40-44y) – 0.595(age 45-49y) - 0.584(age 50-54y) – 0.720(age 55-59y) – 0.676(age 60-64y) – 0.612(age 65-69y) – 0.961(age 70-74y) - 0.565(age 75-79y) - 0.768(age 80-84y) – 1.377(age 85y+) (R^2^=95.1%)

*Notes*: IMD: Index of Multiple Deprivation. Weight values are in kg. Reference groups: age-group (16-17y); IMD (Q1: least deprived); educational status (degree or equivalent); ethnic group (White); smoking status (current cigarette smoker); general health (very good/good).

**Table S6**

**Regression of difference between self-reported and measured BMI on measured BMI by sex**

|  | **Men** | | |  | **Women** | | |
| --- | --- | --- | --- | --- | --- | --- | --- |
|  | **β** | **95% CI** | **P** |  | **β** | **95% CI** | **P** |
| Intercept | 2.19 | 1.90 to 2.49 | <0.001 |  | 1.61 | 1.39 to 1.82 | <0.001 |
| BMI_measured_ | -0.12 | -0.13 to -0.10 | <0.001 |  | -0.10 | -0.11 to -0.09 | <0.001 |
| R^2^ | 13.5% |  |  |  | 12.9% |  |  |

BMI: body mass index; CI: confidence interval. Dependent variable: self-reported minus measured BMI. Men and women with valid self-reported and measured height and weight (split-sample B).

**Regression of difference between corrected (full model) and measured BMI on measured BMI by sex**

|  | **Men** | | |  | **Women** | | |
| --- | --- | --- | --- | --- | --- | --- | --- |
|  | **β** | **95% CI** | **P** |  | **β** | **95% CI** | **P** |
| Intercept | 2.11 | 1.81 to 2.40 | <0.001 |  | 1.84 | 1.63 to 2.05 | <0.001 |
| BMI_measured_ | -0.08 | -0.09 to -0.07 | <0.001 |  | -0.07 | -0.08 to -0.06 | <0.001 |
| R^2^ | 6.4% |  |  |  | 6.6% |  |  |

BMI_M_: body mass index; CI: confidence interval. Dependent variable: corrected minus measured BMI. Men and women with valid self-reported and measured height and weight (split-sample B).

**Table S7**	**Regression of measured height on self-reported height and age by sex**

|  | **β** | **95% Confidence Interval** | **P** |
| --- | --- | --- | --- |
|  | *Men (n=12,376)* | | |
| Intercept | 314.0902 | -9.243173 to 637.4236 | 0.057 |
| Height | -3.755022 | -9.231661 to 1.721617 | 0.179 |
| Height^2^ | 0.0239478 | -0.0069428 to 0.0548384 | 0.129 |
| Height^3^ | -0.0000406 | -0.0000986 to 0.0000175 | 0.171 |
| Age | 0.0787617 | 0.0637293 to 0.0937942 | <0.001 |
| Age^2^ | -0.0012556 | -0.0014043 to -0.0011068 | <0.001 |
| R^2^ | **88.5%** |  |  |
|  | *Women (n=14,659)* | | |
| Intercept | 160.3468 | -162.7323 to 483.426 | 0.331 |
| Height | -1.736024 | -7.65337 to 4.181321 | 0.565 |
| Height^2^ | 0.0159004 | -0.0201807 to 0.0519815 | 0.388 |
| Height^3^ | -0.0000321 | -0.0001054 to 0.0000411 | 0.390 |
| Age | 0.1119681 | 0.0977273 to 0.1262089 | <0.001 |
| Age^2^ | -0.0016046 | -0.0017469 to -0.0014624 | <0.001 |
| R^2^ | **87.0%** |  |  |

*Notes:* Dependent variable: measured height. Men and women with valid self-reported and measured height and weight. Parameters estimated using split-sample A.

_____________________________________________________________________________

*Prediction equations:* These regression parameters can be used to obtain an estimate of the corrected height measurements for participants. Note that age in our analysis was trimmed to a maximum of 90 (range: 16 to 90). Height values are in cm.

**adjusted height (men)** = 314.0902 + (-3.755022 × height) + (0.0239478 × height^2^) + (-0.0000406 × height^3^) + (0.0787617 × age) + (-0.0012556 × age^2^) (R^2^=88.5%)

**adjusted height (women)** = 160.3468 + (-1.736024 × height) + (0.0159004 × height^2^) + (-0.0000321 × height^3^) + (0.1119681 × age) + (-0.0016046 × age^2^) (R^2^=87.0%)

**Table S8**	**Regression of measured weight on self-reported weight and age by sex**

|  | **β** | **95% Confidence Interval** | **P** |
| --- | --- | --- | --- |
|  | *Men (n=12,376)* | | |
| Intercept | 7.2919 | 2.81962 to 11.76418 | 0.001 |
| Weight | 0.7580023 | 0.6139772 to 0.9020274 | <0.001 |
| Weight^2^ | 0.0026345 | 0.0011405 to 0.0041284 | 0.001 |
| Weight^3^ | -8.66e-06 | -0.0000137 to -3.58e-06 | 0.001 |
| Age | 0.0316071 | 0.0073247 to 0.0558896 | 0.011 |
| Age^2^ | -0.0001926 | -0.0004235 to 0.0000382 | 0.102 |
| R^2^ | **94.3%** |  |  |
|  | *Women (n=14,659)* | | |
| Intercept | 1.281195 | -1.83189 to 4.39428 | 0.420 |
| Weight | 0.9487828 | 0.827933 to 1.069633 | <0.001 |
| Weight^2^ | 0.001376 | -0.0001289 to 0.0028809 | 0.073 |
| Weight^3^ | -7.55e-06 | -0.0000136 to -1.50e-06 | 0.014 |
| Age | 0.0083899 | -0.0115418 to 0.0283216 | 0.409 |
| Age^2^ | -0.0000739 | -0.0002655 to 0.0001178 | 0.450 |
| R^2^ | **95.0%** |  |  |

*Notes:* Dependent variable: measured weight. Men and women with valid self-reported and measured height and weight. Parameters estimated using split-sample A.

______________________________________________________________________________________

*Prediction equations:* These regression parameters can be used to obtain an estimate of the corrected weight measurements for participants. Note that age in our analysis was trimmed to a maximum of 90 (range: 16 to 90). Weight values are in kg.

**adjusted weight (men)** = 7.2919 + (0.7580023 × weight) + (0.0026345 × weight^2^) + (-0.00000866 × weight^3^) + (0.0316071 × age) + (-0.0001926 × age^2^) (R^2^=94.3%)

**adjusted weight (women)** = 1.281195 + (0.9487828 × weight) + (0.001376 × weight^2^) + (-0.00000755 × weight^3^) + (0.0083899 × age) + (-0.0000739 × age^2^) (R^2^=95.0%)

**Table S9	Cross-tabulation of measured and corrected BMI categories by sex (split-sample B)**

| **Corrected BMI** | **Measured BMI** | | | | |
| --- | --- | --- | --- | --- | --- |
|  | **Underweight** | **Normal** | **Overweight but not obese** | **Obese I & II** | **Obese III** |
|  | **N (%)** | **N (%)** | **N (%)** | **N (%)** | **N (%)** |
| **Men (n=5,297):** |  |  |  |  |  |
| Underweight | 54 (55.3) | 20 (1.1) | 0 (0.0) | 0 (0.0) | 0 (0.0) |
| Normal | 44 (44.7) | 1565 (83.1) | 228 (9.3) | 1 (0.1) | 0 (0.0) |
| Overweight but not obese | 0 (0.0) | 297 (15.8) | 2048 (84.0) | 211 (15.6) | 0 (0.0) |
| Obese I & II | 0 (0.0) | 0 (0.0) | 162 (6.7) | 1120 (82.8) | 30 (28.1) |
| Obese III | 0 (0.0) | 0 (0.0) | 0 (0.0) | 22 (1.6) | 76 (71.9) |
| Sensitivity | 55% | 83% | 84% | 83% | 72% |
| Specificity | 100% | 93% | 85% | 96% | 100% |
| κ | 0.74 |  |  |  |  |
| **Women (n=6,145):** |  |  |  |  |  |
| Underweight | 93 (66.7) | 36 (1.5) | 0 (0.0) | 0 (0.0) | 0 (0.0) |
| Normal | 46 (33.3) | 2115 (88.1) | 212 (11.9) | 4 (0.3) | 0 (0.0) |
| Overweight but not obese | 0 (0.0) | 247 (10.3) | 1430 (80.5) | 183 (15.6) | 0 (0.0) |
| Obese I & II | 0 (0.0) | 2 (0.1) | 134 (7.6) | 967 (82.2) | 48 (27.5) |
| Obese III | 0 (0.0) | 0 (0.0) | 0 (0.0) | 23 (1.9) | 127 (72.6) |
| Sensitivity | 67% | 88% | 80% | 82% | 73% |
| Specificity | 99% | 92% | 89% | 96% | 100% |
| κ | 0.76 |  |  |  |  |

*Notes:* Cell counts are weighted (rounded); estimates are column percentages. BMI: body mass index. Underweight: BMI <18.5kg/m^2^; Normal: ≥18.5 – 24.9kg/m^2^; Overweight but not obese: ≥25.0 – 29.9kg/m^2^; Obese I & II: ≥30.0 – 39.9kg/m^2^; Obese III: ≥40.0kg/m^2^. Participants with valid self-reported and measured height and weight (n=11,442: split-sample B). κ (Cohen’s kappa).

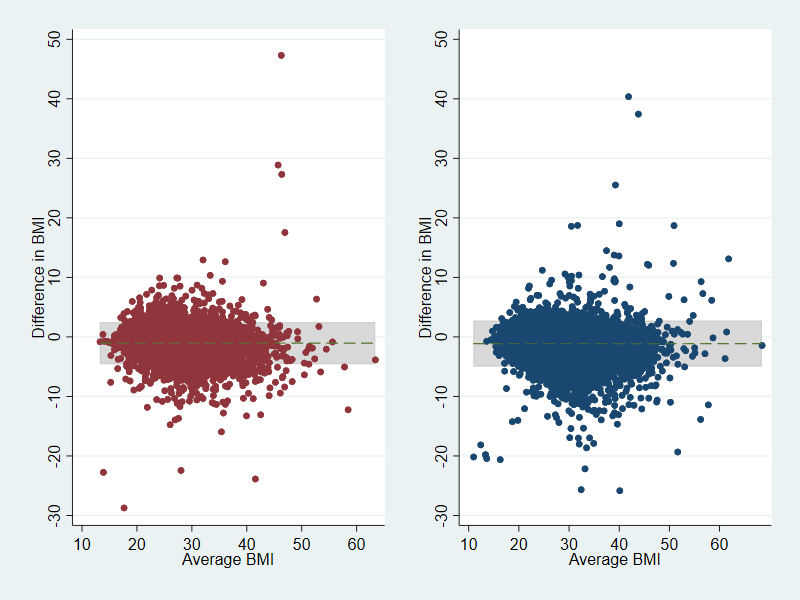

***Figure S1****:* Bland-Altman plot of the difference between self-reported and measured BMI vs. the average of these two measures of BMI (men: left-panel; women : right-panel). Plots produced using unweighted data from n=38,942 participants with valid self-report and measured height and weight (includes participants with an absolute difference (self-reported minus measured) in height or weight ≥4SD from the mean).

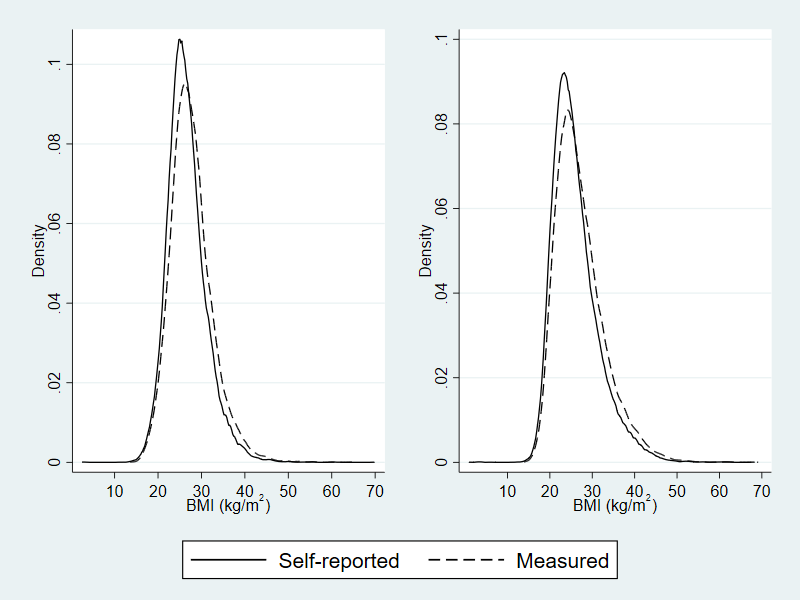

***Figure S2****: Distributions for self-reported and measured BMI* (men: left-panel; women : right-panel). Plots produced using data from n=38,942 participants with valid self-report and measured height and weight (*includes* participants with an absolute difference (self-reported minus measured) in height or weight ≥4SD from the mean).

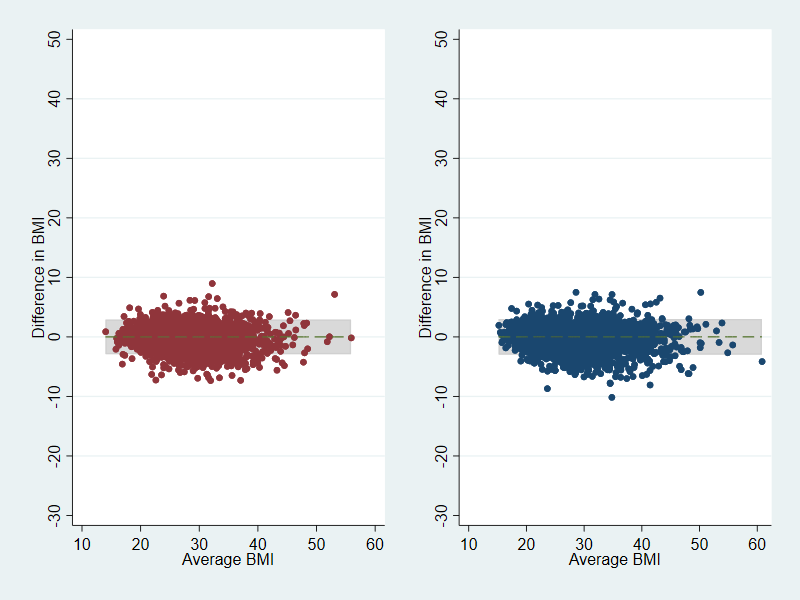

***Figure S3:*** Bland-Altman plot of the difference between corrected and measured BMI vs. the average of these two measures of BMI (men: left-panel; women : right-panel). Plots produced using data from n=11,442 participants with valid self-report and measured height and weight (split-sample B).

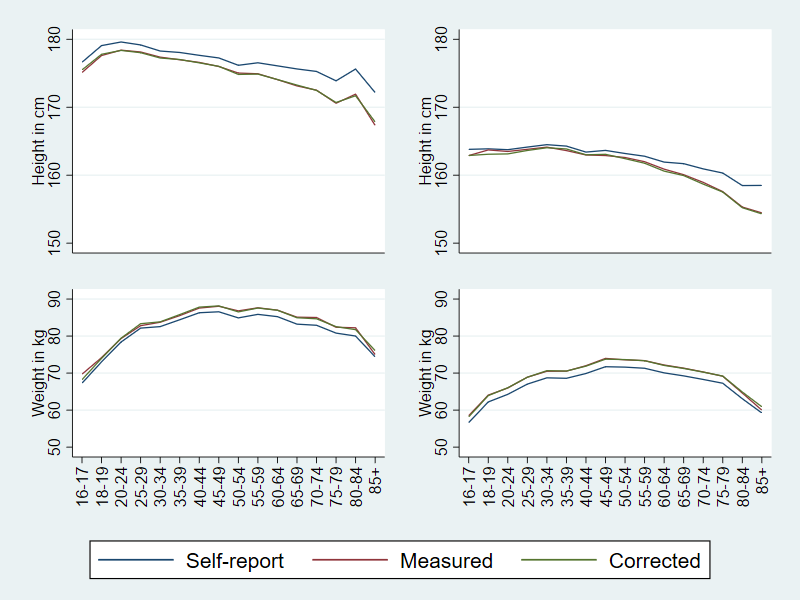

***Figure S4*** Self-reported, measured and corrected height and weight by age-group and sex (men: left-panel; women : right-panel).
